# Functional organization of multimorbidity links chronic disease incidence to shared genetic architecture

**DOI:** 10.64898/2026.10.02.26364618

**Authors:** Daniel Igumnov, Ekaterina Melnikova, Semyon Matrenok, Kirill Denisov, Klim Kostiuk, Arina Nikolskaya, Liza Lopatina, Salih Tuna, Konstantin Avchaciov, Amir Feizi, Peter Fedichev

## Abstract

The multistage model of carcinogenesis explains the steep rise of cancer incidence with age through accumulated events, and reliability theory extends this principle to systems whose failure requires several components. We asked whether the same idea could explain why chronic diseases differ in how rapidly incidence rises with age, and whether multimorbidity could independently reveal the processes involved. In UK Biobank, linked health records allowed pre-assessment disease co-occurrence and subsequent incidence to be analysed separately. Using generative representation learning, we derived 16 functional units (FUs) from co-occurrence patterns across 232 diagnoses in 502,355 participants. For each diagnosis, we counted linked FUs and compared this breadth with subsequent incidence. Diagnoses linked to more FUs showed steeper age dependence of incidence (Spearman ***ρ* = 0.50**; 0.46 among 196 diagnoses with rising hazards), and this relationship persisted in an age-free representation. The FUs had distinct genetic architectures, with lead-variant directions preserved in FinnGen and the Million Veteran Program. Their GWAS reached 34 regions missed by diagnosis and BMI comparators, while fine-mapping and molecular evidence connected FU loci to candidate genes enriched for approved drug targets. Together, these results extend a multistage principle from cancer to chronic disease and show that multimorbidity structure explains variation in age-dependent incidence.

---

Chronic diseases differ strikingly in how their incidence changes with age. The multistage model of carcinogenesis explains the steep rise of cancer incidence with age as the accumulation of multiple events [1], and the same logic has been applied to individual neurodegenerative diseases, including amyotrophic lateral sclerosis [2]. Reliability theory generalizes this principle to systems whose failure depends on several components [3]. A complementary threshold model explains the rise of age-related diseases through a shared accumulating variable with disease-specific thresholds [4]. These frameworks differ mechanistically, but they share an important limitation: the disease-specific event count, component number or threshold is inferred from the incidence curve it is meant to explain. Whether the relevant disease complexity can be measured independently has therefore remained unresolved.

We reasoned that multimorbidity could provide this independent measurement. If chronic diseases arise through combinations of shared physiological processes, diseases involving more of them should be embedded more broadly across recurring patterns of disease co-occurrence. This gives a direct hypothesis: multimorbidity breadth, measured without using age at disease onset, should explain variation in how rapidly subsequent incidence rises with age. Existing clinical summaries do not directly measure this overlapping structure. Frailty indices [5] and biological-age measures [6, 7] compress health into global scores, whereas organ-specific ageing models [8] partition ageing by anatomical system. Large biobanks now provide the linked longitudinal records needed to separate disease co-occurrence before assessment from subsequent incidence, while also providing molecular and genetic measurements with which to test the biological structure of any inferred dimensions [9].

Human genetics provides an independent test of whether such dimensions capture shared disease biology rather than arbitrary statistical structure. If a functional dimension represents a recurring physiological process, it should have a coherent heritable architecture, and related diagnoses should contribute concordant genetic effects. Previous topic models of diagnosis co-occurrence [10], linear distillation of biobank traits [11] and longitudinal disease-signature models [12] have shown that joint phenotypes can recover genetic associations beyond individual input phenotypes. We therefore use genetics as a second readout of the multimorbidity organization: distinct FU architectures test biological separation, reconstruction from matched diagnoses in FinnGen and the Million Veteran Program (MVP) tests portability of the shared genetic direction, and comparison with diagnosis and body-mass-index (BMI) GWAS tests where aggregation concentrates signal that remains below genome-wide significance in conventional phenotypes.

Here we test this framework in 502,355 UK Biobank participants, where linked health records allow pre-assessment multimorbidity and subsequent disease incidence to be analysed separately. Using generative representation learning on diagnoses accumulated before assessment, we derive 16 continuous functional units (FUs) that capture recurring multimorbidity patterns across 232 chronic diagnoses (Fig. 1). We then count how many FUs are linked to each diagnosis and compare this breadth with the independently fitted age dependence of subsequent incidence. Age at disease onset and genotype do not enter representation learning, and we repeat the analysis after removing participant age from the model. We next test whether the same FUs have distinct molecular and genetic architectures, whether their genetic directions are preserved in FinnGen and MVP, and whether FU GWAS concentrate distributed genetic signal across diagnosis boundaries. Finally, fine-mapping and molecular evidence connect FU loci to candidate genes and established therapeutic biology.

**Fig. 1.**
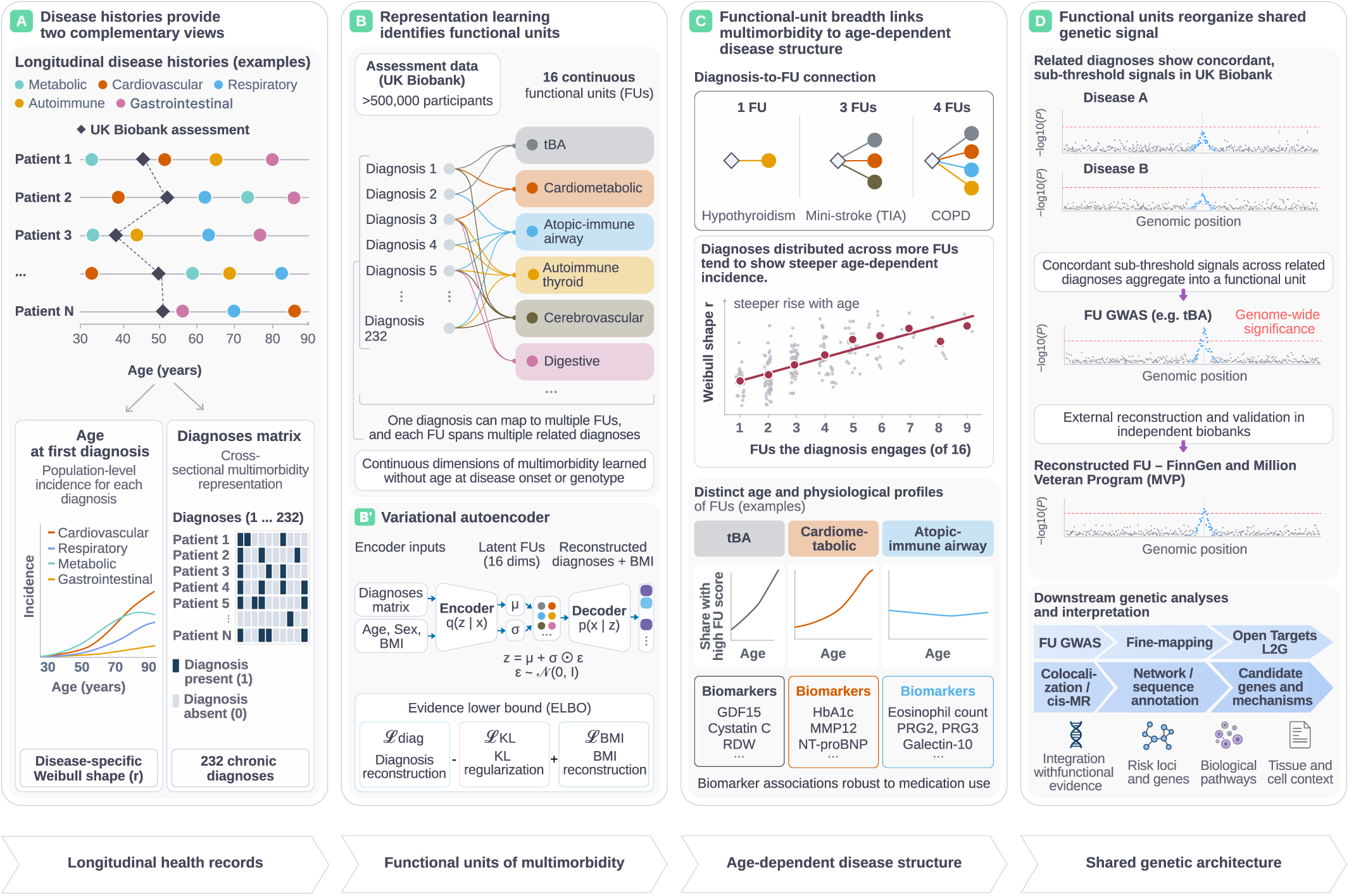
Functional units (FUs) provide an independent measure of multimorbidity breadth for testing variation in age-dependent disease incidence. **a,** Longitudinal diagnosis histories provide two views of the same records. First recorded diagnoses define age-specific incidence after assessment, while diagnoses present at assessment form the cross-sectional matrix used for representation learning. **b,** Generative representation learning of 232 chronic diagnoses, together with BMI, yields 16 continuous FUs capturing recurring multimorbidity patterns. The diagnosis-to-FU map is many-to-many, so one diagnosis can engage several FUs and one FU can span diagnoses across clinical domains. **b*^′^*,** Variational-autoencoder implementation. Age at assessment and sex enter the encoder, sex enters the decoder, and 16 of 32 latent coordinates are retained by the mean Kullback– Leibler activity threshold. Exact age at first recorded diagnosis and genotype are not representation inputs. **c,** FU breadth is the number of FUs linked to a diagnosis. The central test is whether diagnoses engaging more FUs show faster of acceleration incidence with age. Lower panels show age profiles and example biomarkers of three FUs. **d,** FU genome-wide association scans test whether the same dimensions carry coherent shared genetic signal. Lead variants are evaluated by diagnosis-weighted reconstruction in external biobanks and followed by fine-mapping, molecular evidence and therapeutic annotation. Panels summarize the study design; quantitative results are shown in Figs. 2–5.

## 1.1 Multimorbidity breadth explains variation in incidence acceleration with age

Across 232 chronic diagnoses, first-diagnosis incidence between ages 44 and 76 varied markedly in both magnitude and age dependence (Fig. 2a). Nine cancer tissue groups provide a reference for the steep age dependence of multistage carcinogenesis without implying a shared somatic mechanism. We summarized each chronic-disease curve by the age-scaling parameter *r* in the cause-specific hazard *h*(*t*) = *at^r−^*^1^, where *t* is attained age and *a >* 0 sets the overall hazard scale, fitted with left truncation at cohort entry (§3.3). For *r >* 0, this is also the Weibull shape parameter; *r −* 1 is the slope of log hazard on log age and quantifies how rapidly the hazard accelerates with age on this scale. The fitted *r* ranged from *−*2.05 to 11.0 (median 3.13). For 196 diagnoses *r ≥* 1, the fitted hazard rose with age, and 183 had a 95% interval lying entirely above 1. The other 36 diagnoses had flat or declining fitted hazards over the observed ages and were mainly conditions commonly recorded before recruitment. We therefore analyse all 232 diagnoses and, separately, the 196 with rising hazards, the range to which the reliability-theory interpretation applies.

**Fig. 2.**
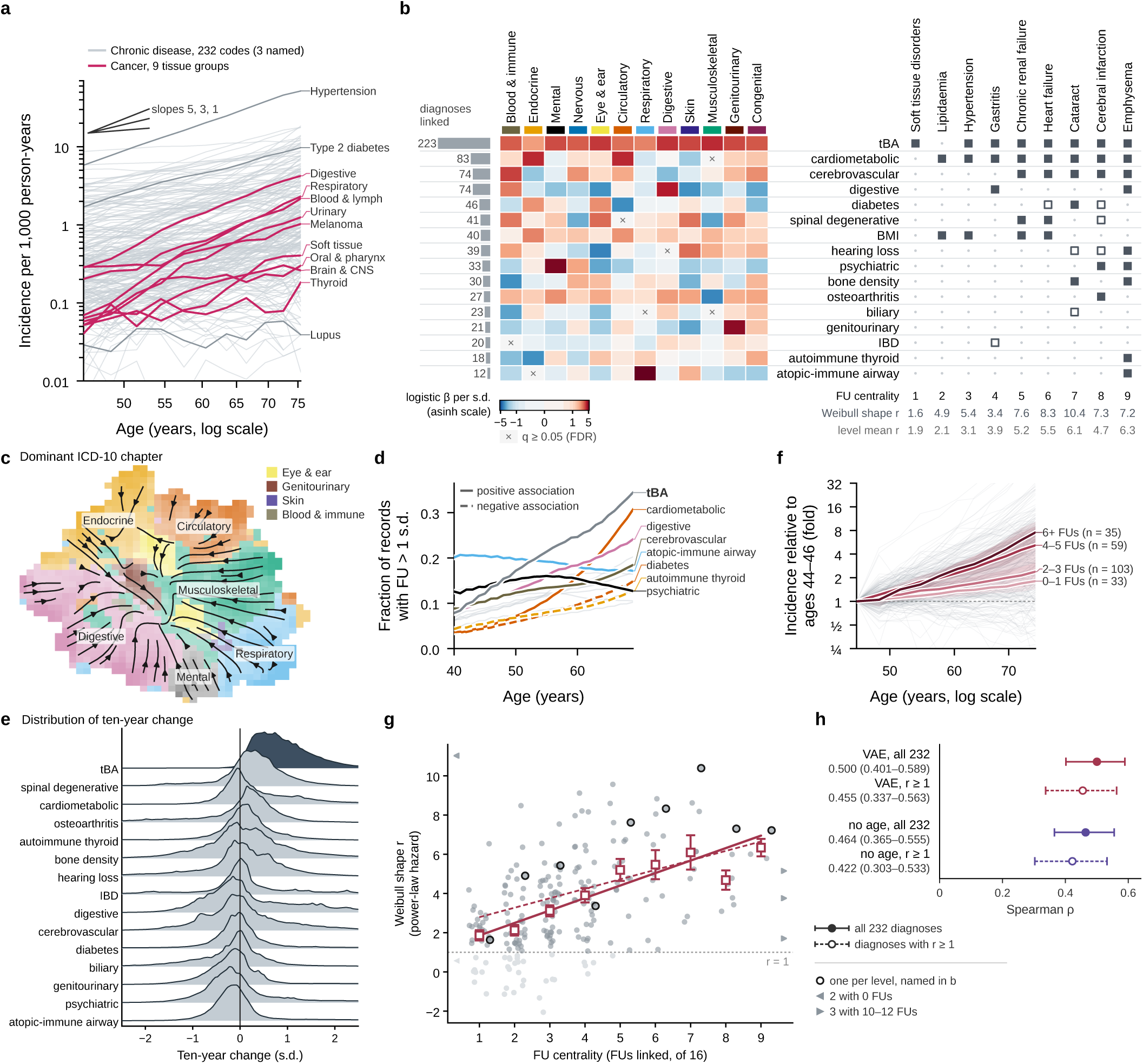
Diagnoses linked to more functional units (FUs) show faster acceleration of incidence with age. **a**, First-diagnosis incidence for 232 chronic diagnoses and nine cancer groups. **b**, Left, associations between FU scores and ICD-10 chapters (*N* = 498,788), with the number of diagnoses linked to each FU. Right, FU memberships for one representative diagnosis at each centrality level. **c**, UMAP of FU scores for 293,061 participants with at least two diagnoses, coloured by the most common dominant ICD-10 chapter in each cell. Streamlines show average projected ten-year displacements for illustration and are not trajectories. **d**, Fraction of records with each FU above 1 s.d. by attained age. **e**, Ten-year change in each disease FU among 147,839 participants younger than 60 years at assessment who acquired a new diagnosis. **f**, Chronic-disease incidence relative to ages 44–46, grouped by FU centrality. Lines are geometric means and ribbons are 95% chapter-bootstrap intervals. **g**, age-scaling parameter *r* of the hazard *h*(*t*) = *at^r−^*^1^ against FU centrality. Light grey points have *r <* 1. Lines are Theil–Sen fits and squares are level means *±* s.e.m. **h**, Spearman *ρ* between centrality and *r* for the primary VAE and a model retrained without age input. Bars are 95% within-chapter bootstrap intervals over diagnoses at the fixed penalty and do not include model-training uncertainty. IBD, inflammatory bowel disease. tBA, thermodynamic biological age.

To derive shared disease structure independently from the incidence curves, we applied generative representation learning to diagnoses accumulated before assessment and BMI in 502,355 participants (Fig. 2b). The implementation used a variational autoencoder with age at assessment and sex available to the encoder (Fig. 1 and §3.4). The latent dimensions were learned without expert-defined disease groupings, genotype or age at first diagnosis. Descriptive names were assigned only after fitting, from their diagnosis and ICD-10 chapter association profiles with genetic, biomarker and medication results consulted. Sixteen of 32 latent coordinates had mean marginal Kullback–Leibler activity greater than 0.05 and were retained as FUs. Each participant therefore has a continuous score on each FU, describing position along a learned multimorbidity dimension rather than the probability of any single disease; diagnoses can associate with an FU in either direction. The FU profiles span cardiometabolic, cerebrovascular, gastrointestinal, musculoskeletal, psychiatric, immune and endocrine patterns, with some concentrated within one clinical domain and others crossing several. One of the 16 dimensions, the broadest, was linked to 223 of the 232 diagnoses and is termed thermodynamic biological age (tBA). The name follows Tarkhov *et al.* [13], in which tBA summarizes accumulated transitions among metastable functional states. Here it denotes the learned broad disease-burden dimension and is not assumed to be identical to the theoretical entropy variable.

The continuous FU scores varied systematically across participants and shifted within individuals as diagnoses accumulated. The participant map shows structured variation in the dominant ICD-10 chapter of participants’ diagnoses (Fig. 2c), and tBA broadly follows prevalent diagnosis burden while retaining distinct structure (Extended Data Fig. 1). Between ages 41 and 69, the fraction of records above 1 s.d. rose most for the cardiometabolic and tBA FUs and fell for the atopic–immune airway FU (Fig. 2d). Over ten years, the tBA FU decreased in only 1.2% of 147,839 participants who acquired a new diagnosis, whereas the other disease FUs decreased in 28% to 65% (Fig. 2e and §3.5).

We then tested the central prediction: diagnoses linked to more FUs should show a steeper rise in incidence with age than diagnoses linked to only a few. We quantified multimorbidity breadth as FU centrality, the number of FUs linked to each diagnosis, and age dependence by the separately fitted parameter *r*. For each FU–diagnosis pair we fitted an *L*_1_-penalized logistic model of diagnosis status on one FU score, age and sex. For association selection, the target diagnosis was removed from the encoder input before its FU score was computed, yielding 804 retained FU–diagnosis links at the analysis penalty. FU centrality therefore quantifies how broadly a diagnosis is embedded across the learned multimorbidity dimensions. Crucially, centrality and incidence acceleration are estimated from non-overlapping disease events: centrality uses diagnoses recorded before assessment, whereas *r* uses first diagnoses recorded after assessment, so no diagnosis event enters both quantities.

The relationship is visible without a hazard model. Incidence curves normalized to ages 44–46 become progressively steeper as diagnoses engage more FUs (Fig. 2f). Across all 232 diagnoses, the age-scaling parameter *r* increased with FU centrality (Spearman *ρ* = 0.500, 95% confidence interval (CI) 0.401–0.589), and the association remained among the 196 diagnoses with rising hazards (*ρ* = 0.455, 95% CI 0.337–0.563, Fig. 2g,h). We accounted for choosing the *L*_1_ penalty by repeating the selection across the full penalty path within 100,000 chapter-stratified permutations. None reached the observed association in either diagnosis set (*P <* 10*^−^*^5^, §3.3). The relationship also held in diagnoses not used to choose the penalty. Choosing the penalty on one random half and evaluating it on the other gave median held-out correlations of 0.466 for the 232-diagnosis set and 0.407 for the 196-diagnosis set.

The gradient extends through the central range of the data rather than arising only from the 36 diagnoses with flat or declining hazards. Among the 196 diagnoses with *r ≥* 1, mean *r* rose from 2.50 at one linked FU to 6.09 at seven. The 36 diagnoses with *r <* 1 were linked to fewer FUs on average, with means of 2.19 for *r ≤* 0 and 2.20 for 0 *< r <* 1, compared with 3.70 among the 196 (Fig. 2g). Above seven FUs each centrality level contains eight diagnoses or fewer and its mean becomes correspondingly unstable. The monotonic rise across the central range is consistent with the composition hypothesis: diseases embedded across more functional dimensions show faster acceleration of incidence with age. We do not interpret the numerical increment per FU as a parameter-free prediction.

Because age at assessment entered the primary encoder, FU scores could carry age information and the centrality association might simply reflect age-related diagnoses loading on age-related dimensions. To test this, we retrained the same model without age at assessment, allowing the number of active dimensions to be selected again by the same activity rule. Fifteen dimensions were retained, and most primary FUs had closely corresponding age-free dimensions (Extended Data Fig. 2b). At the fixed penalty used for the primary analysis, the positive centrality–*r* relationship remained across all 232 diagnoses (Spearman *ρ* = 0.464, 95% CI 0.365–0.555) and among the 196 with rising hazards (*ρ* = 0.422, 0.303–0.533, Fig. 2h). Path-adjusted permutation gave *P <* 10*^−^*^5^ and *P* = 2 *×* 10*^−^*^5^, respectively.

The relationship was also stronger for the nonlinear FUs than for two linear decompositions of the same diagnosis matrix, logistic principal-components analysis and logistic singular-value decomposition, which compress the same diagnosis data without the nonlinear encoder. On the common 100,471-participant association split, logistic principal-components analysis gave *ρ* = 0.263 across all 232 diagnoses and 0.294 among the 196, while logistic singular-value decomposition gave 0.274 and 0.282, respectively (Extended Data Fig. 2c–e and §3.8). Split-half penalty transfer, in which the penalty selected on one diagnosis half was evaluated on the other, showed the same ordering. Neither age at first recorded diagnosis nor any fitted incidence parameter entered representation learning. Together with the no-age retraining, these analyses show that the centrality–*r* relationship is not a direct consequence of supplying age to the model or of a linear reduction of the diagnosis matrix to fewer dimensions.

## 1.2 Functional units have distinct and portable biological architectures

Having established that FU breadth tracks incidence acceleration, we next asked whether the FUs capture biologically distinct dimensions rather than arbitrary statistical structure. FU profiles carried prognostic information beyond diagnosis count. A cross-validated mortality model using the 15 disease FUs, age and sex reached Harrell’s *C* = 0.740, compared with 0.723 for age, sex and diagnosis count, and diagnosis count added only 0.0004 (95% CI 0.0002–0.0006) once the FU risk score was present (§3.7). The largest mortality-aligned hazard ratios per standard deviation were 1.32 (95% CI 1.30–1.33) for the cardiometabolic FU, 1.19 for cerebrovascular and 1.16 for psychiatric. These estimates describe the information captured by the FU profile. Their signs are not an independent validation because FU orientation used mortality during naming.

The FUs also map onto distinct molecular profiles measured at assessment. Glucose and glycated haemoglobin rise with the cardiometabolic FU, whereas eosinophil-related measurements mark the atopic–immune airway FU (Extended Data Fig. 3a). Medication use helps explain part of these cross-sectional associations. Participants at the disease end of the diabetes and atopic–immune airway FUs used metformin and inhaled corticosteroids 61-fold and 17-fold more often than age- and sex-matched participants outside that decile (Extended Data Fig. 3b). Single medication classes accounted for more than half of eight of the 27 displayed molecular associations but less than one quarter of each airway-FU eosinophil association (§3.6).

Genome-wide association established that the FUs also have distinct common-variant architectures. In 423,955 European-ancestry participants, all 16 FU scores had detectable SNP heritability with *h*^2^*/*s.e*. ≥* 5.35 and together yielded 979 genome-wide-significant leads over the 16 scans (Fig. 3a,b and Extended Data Fig. 4). Genetic correlations between FUs were low. Across the 120 FU pairs, the median absolute *r_g_* was 0.097, lower than in all 2,000 random rotations of the FU basis, and only the digestive–tBA pair exceeded 0.5 (*r_g_* = 0.54, Fig. 3c and Extended Data Fig. 5b). This separation relative to random rotations supports genetically distinct dimensions rather than an arbitrary rotation of a common genetic axis. FU–diagnosis genetic correlations also tracked the clinical organization. Among 384 associations passing a 5% false-discovery rate, 13 of the 14 FUs named for an ICD-10 chapter had more significant correlations in their named chapter than in any other (Extended Data Fig. 5a,c).

**Fig. 3.**
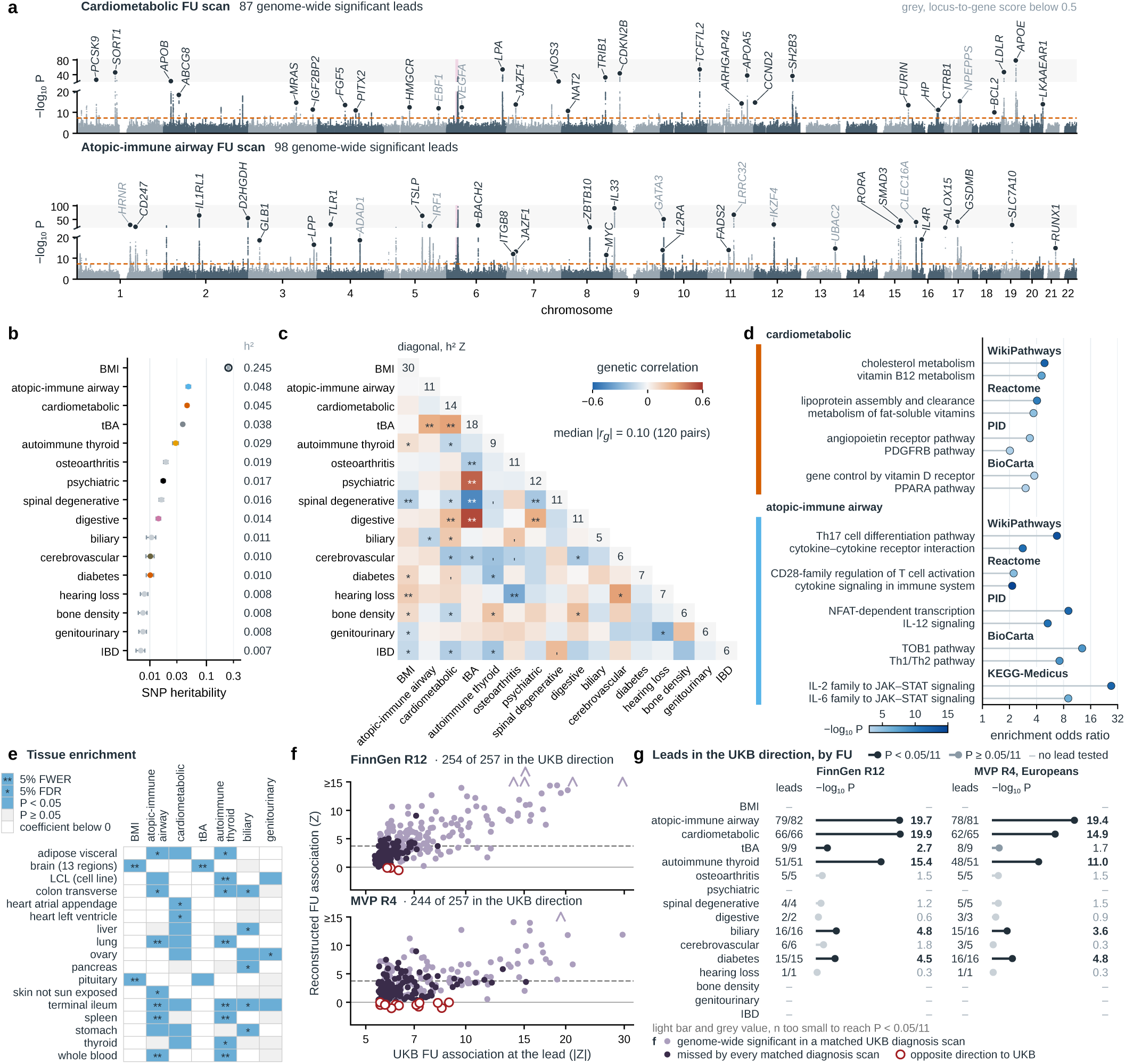
Functional units (FUs) have distinct genetic architectures whose lead-variant directions are preserved in external cohorts. **a**, GWAS of the cardiometabolic and atopic–immune airway FUs in 423,955 UK Biobank participants. Dashed line, *P* = 5 *×* 10*^−^*^8^. Pink band, major histocompatibility complex. Labels are the top gene of the nearest locus-to-gene credible set within 500 kb. **b**, Observed-scale SNP heritability *±* s.e. for all FUs. **c**, Pairwise genetic correlations. The diagonal shows *h*^2^*/*s.e. and symbols mark nominal, false-discovery-rate and Bonferroni significance across 120 pairs. **d**, Representative pathway enrichments for the cardiometabolic and atopic–immune airway FUs. **e**, GTEx tissue enrichments for FUs reaching 5 % false-discovery rate across 784 FU–tissue tests. The brain row shows the strongest significance level reached by any of its 13 GTEx regions. **f**, UK Biobank FU lead variants tested through diagnosis-weighted composites in FinnGen and the Million veteran Program (MVP). Positive reconstructed *Z* indicates agreement with the UK Biobank direction. **g**, Directional agreement by FU with one-sided exact binomial tests against 0.5. IBD, inflammatory bowel disease. tBA, thermodynamic biological age. LCL, lymphoblastoid cell line.

Pathway and tissue enrichments further separated the FU architectures. The cardiometabolic FU was enriched for lipoprotein and sterol processes, whereas the atopic–immune airway FU was enriched for adaptive immune and cytokine pathways (Fig. 3d). At a 5% family-wise error rate across 784 FU–tissue tests, the BMI and tBA FUs were enriched in brain and the autoimmune-thyroid and atopic–immune airway FUs in immune tissues or lung (Fig. 3e). The brain enrichment of the tBA FU is usual for GWAS of BMI, educational attainment and psychiatric traits [14], so health behaviour and healthcare contact are candidate sources beside disease biology. At a 5% false-discovery rate, cardiometabolic also reached heart, biliary reached liver, genitourinary reached ovary and autoimmune thyroid reached thyroid. These enrichments support biological coherence of the learned dimensions while remaining downstream consequences of phenotypes learned from the same UK Biobank diagnosis records.

The distributed genetic signal carried by the FUs is directionally portable beyond UK Biobank. For eleven FUs, we formed in each external cohort a composite of matched diagnosis-level associations using chromosome-held-out weights learned in UK Biobank (§3.13). This reconstructed composite is not a second measurement of the FU, which is available only in UK Biobank. We define a lead variant as the most significant variant at a genome-wide-significant FU locus. Of 257 lead variants tested in each cohort, 250 of them in both, the reconstructed composites preserved the UK Biobank FU direction at 254 in FinnGen (98.8%) and 244 in MVP Europeans (94.9%), compared with 52.4% and 57.4% at matched background variants (Fig. 3f,g). Agreeing lead variants outnumbered disagreeing lead variants on every chromosome in both cohorts (chromosome-block exact *P* = 2.4 *×* 10*^−^*^7^ in each). In the exploratory China Kadoorie Biobank analysis, 130 of 181 (71.8%) tested lead variants agreed in direction, compared with 51.6% at background variants (Extended Data Fig. 6c,d).

The same pattern held at lead variants where no matched constituent diagnosis reached genome-wide significance in UK Biobank. Among these variants, the reconstructed composite agreed with the FU direction at 78 of 80 in FinnGen and 134 of 140 in MVP (Extended Data Fig. 6e,f). Within UK Biobank, FU effects also showed excess directional agreement in five smaller non-European genetic-ancestry groups, although those groups were not powered for genome-wide replication (Extended Data Fig. 6a,b). Thus the cross-disease genetic pattern identified by the FUs is not confined to a single genome-wide-significant diagnosis or to the discovery sample.

## 1.3 Functional units concentrate genetic signal across diagnoses

The distinct FU architectures suggested that combining diagnoses within an FU could concentrate genetic effects that are individually weak in conventional endpoints. Across 1,716 genomic regions defined jointly from genome-wide-significant variants in the FU, diagnosis, BMI and screened Pan-UK Biobank GWAS (§3.14), the 16 FU GWAS reached 1,102 (Fig. 4a). Excluding regions in which any of the following also reached genome-wide significance—this study’s 232 diagnosis GWAS, the Pan-UK Biobank BMI GWAS [15], 228 matched Pan-UK Biobank diagnosis GWAS or the GIANT–UK Biobank BMI meta-analysis [16]—left 34 regions reached by none of these specified comparators. Nineteen were reached by a disease FU and 15 by the BMI FU alone (Fig. 4b and Extended Data Fig. 7). This is a set difference relative to the stated comparators rather than a claim of universal locus gain. Within that scope, the disease-FU regions are candidate sites where aggregating related diagnoses concentrates shared signal that remains below threshold in the individual GWAS.

**Fig. 4.**
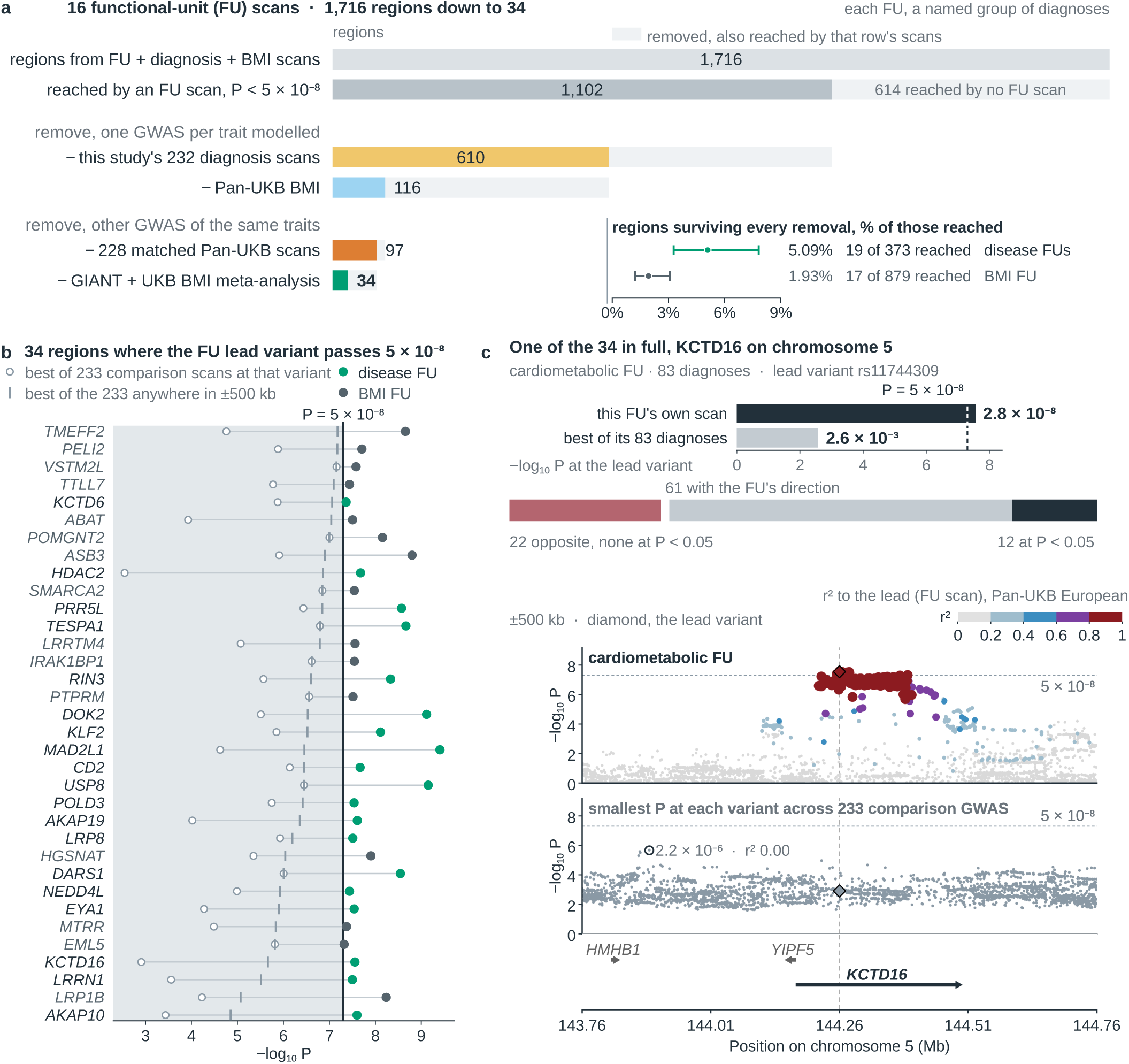
Functional-unit (FU) GWAS reach 34 genomic regions missed by the specified diagnosis and body-mass-index (BMI) comparators. **a**, Starting from 1,716 shared genomic regions, bars show the FU-reached regions remaining after successively removing those also reached by the named comparator scans. The final 34 are independent of removal order. inset, retained regions as a fraction of all regions reached by disease FUs or the BMI FU. Two of the 34 are reached by both a disease FU and the BMI FU and count in both inset rows. **b**, Association strength for the 34 retained regions. Filled points show each FU GWAS at its strongest variant within *±*500 kb of the region centre. Open points show the strongest of this study’s 232 diagnosis GWAS and the Pan-UK Biobank BMI GWAS at that variant and anywhere within *±*500 kb. Gene names are positional labels. **c**, The chromosome 5 region labelled *KCTD16*. Top, the cardiometabolic FU and strongest encoded-diagnosis association at rs11744309. Middle, direction and significance of the 83 encoded diagnoses at the lead. Bottom, regional FU association and the minimum *P* across the 233 comparison GWAS.

The region labelled *KCTD16*, the nearest protein-coding gene to the regional lead variant and therefore a positional label rather than an assigned effector gene, illustrates this distributed pattern (Fig. 4c). The cardiometabolic FU reached genome-wide significance at rs11744309, while the strongest of the 233 diagnosis and BMI GWAS within 500 kb reached *P* = 2.2 *×* 10*^−^*^6^ at an unlinked variant 369 kb away. At the FU lead variant, 61 of the 83 diagnoses encoded by the cardiometabolic FU had a direction of effect aligned with the FU, and all 12 diagnoses with *P <* 0.05 were aligned. The example therefore shows how multiple individually weak but directionally coherent associations can combine into a genome-wide-significant FU signal.

The disease-FU regions also carry support outside the discovery comparison. Eighteen of the 19 belonged to FUs tested in external cohorts, and their available nominations agreed in direction at 14 of 15 tested lead variants in FinnGen, 14 of 15 in MVP and 8 of 12 in CKB. For the cardiometabolic and atopic–immune airway FUs, the GWAS Catalog and GWAS Atlas contained 88 associations at eight of nine corresponding lead variants or high-linkage-disequilibrium proxies, 85 of them at *P ≤* 5 *×* 10*^−^*^8^ (Extended Data Fig. 8a,b). These catalogue records provide a lower bound on external trait support at the loci but do not replicate the FU phenotype or its association. At the atopic–immune airway lead variants, 17 of 20 disease associations involved asthma, allergic rhinitis or atopic dermatitis, consistent with the clinical profile encoded by that FU.

Genetic coherence across the FU–diagnosis links provides a second view of the same distributed architecture. We tested the 804 retained links identified in the centrality analysis above with MR-APSS [17], which models correlated pleiotropy and sample structure in overlapping data. At a 5% false-discovery rate, 126 associations were detected, including 94 links across nine disease FUs and 32 BMI positive-control links. All 126 had the direction of the corresponding mortality-aligned *L*_1_ coefficient, and MR-Egger and weighted mode agreed in direction for 69 and 93 of the 94 disease-FU associations, respectively (Extended Data Fig. 9). Because the diagnoses help define the FUs and exposure and outcome GWAS share participants, these results do not imply that an FU causally affects its constituent diseases. They instead show that the genetic architecture of an FU is distributed across diagnoses within its clinical profile.

## 1.4 Functional-unit loci converge on molecular and therapeutic biology

To connect the shared genetic architecture to molecular mechanisms, we next asked whether FU genetic signal converges on genes with *cis* molecular support. Across the 15 disease FUs, 25 FU–gene pairs involving 21 genes in seven FUs met two criteria in the same plasma-protein assay: a significant MR-Link-2 estimate, providing directional *cis*-genetic evidence relating protein abundance to the FU, and a colocalization posterior of at least 0.8, supporting a shared causal signal (Fig. 5a and §3.17). Twenty-one pairs were supported in UK Biobank plasma proteomics, one in deCODE and three in both. Among the 16 genes whose protein products were measured in both panels, 18 of 26 directed cross-panel comparisons replicated the lead instrument’s protein association in the other panel. UK Biobank protein measurements share participants with the FU GWAS, whereas deCODE provides an independent molecular readout.

**Fig. 5.**
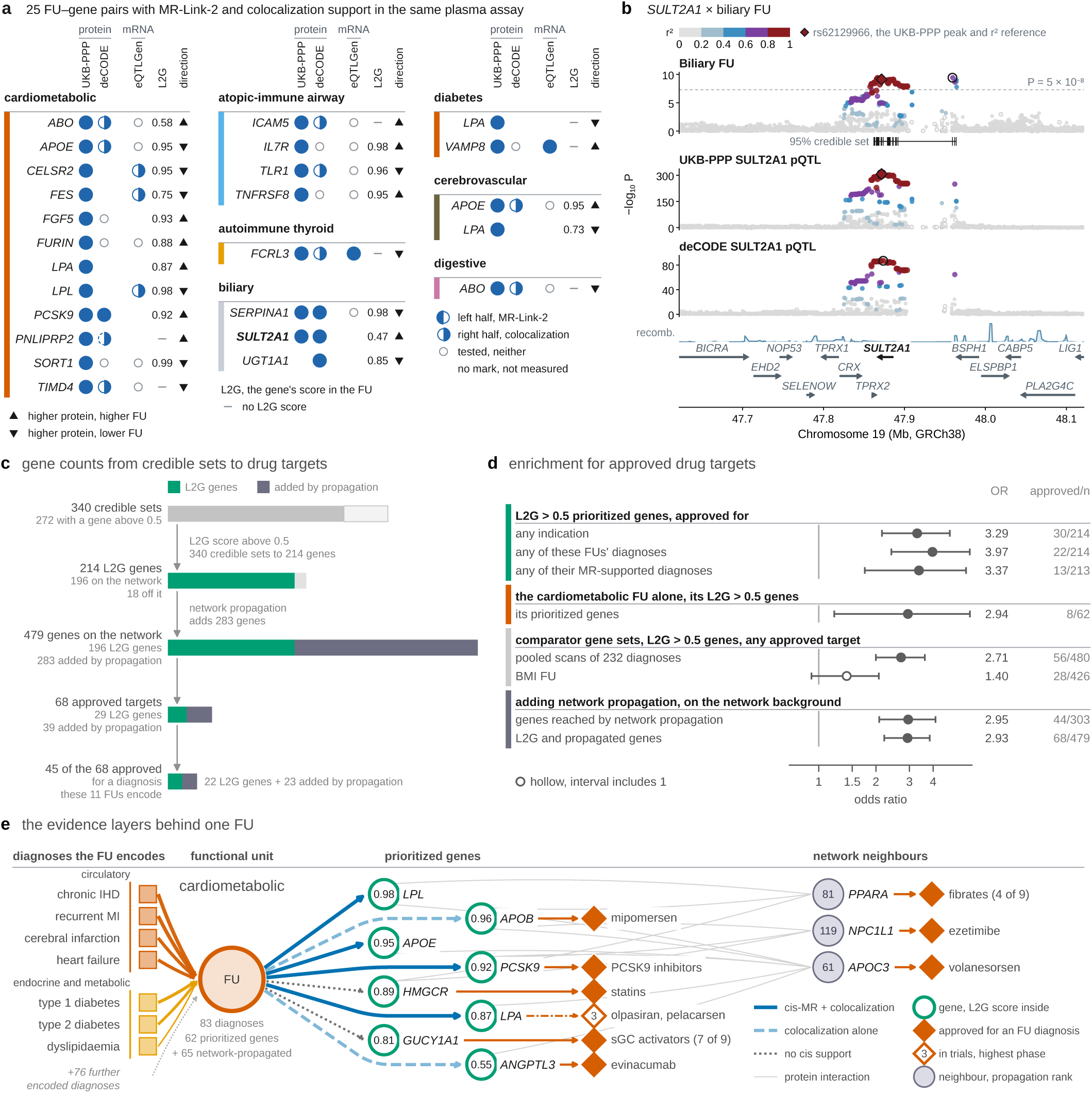
Functional-unit (FU) loci converge on *cis* molecular evidence and approved drug targets. **a**, Twenty-five FU–gene pairs supported by both MR-Link-2 (Benjamini–Hochberg *q <* 0.05 within each FU and cohort) and colocalization (posterior at least 0.8) in the same plasma-protein assay. Half-filled symbols distinguish MR and colocalization support, and columns show UK Biobank, deCODE, eQTLGen and locus-to-gene (L2G) evidence. **b**, Biliary-FU and *SULT2A1* protein associations in UK Biobank and deCODE across the same chromosome 19 region. Open rings mark each track’s strongest variant, and diamonds mark rs62129966, the UKB-PPP peak and *r*^2^ reference. **c**, Flow from 340 fine-mapped credible sets in eleven disease FUs to L2G-prioritized genes, network-propagated genes and approved targets. Propagation added genes for the five of these FUs with more than ten network seeds. **d**, Approved-target enrichment for prioritized and propagated gene sets. points are odds ratios with 95% confidence intervals. **e**, Cardiometabolic-FU evidence map linking selected encoded diagnoses to prioritized genes, approved drugs and network neighbours. Line width shows the mortality-aligned per-pair *L*_1_ coefficient, which is shrunk and carries no standard error. The display rule, support by a *cis* readout or by an approved drug for an encoded diagnosis, was applied to 20 of the FU’s 127 prioritized and propagated genes, and 11 of the 15 that meet it are shown. IHD, ischaemic heart disease. MI, myocardial infarction. sGC, soluble guanylate cyclase.

*SULT2A1* provides a locus-level example in the biliary FU (Fig. 5b). The biliary-FU GWAS peak lay 88 kb from the lead *SULT2A1* protein quantitative-trait locus (pQTL) variant in UK Biobank, and the 50-variant SuSiE credible set for the FU association contained both the FU peak and the pQTL lead. MR-Link-2 passed in UK Biobank (*q* = 5.1 *×* 10*^−^*^5^) and deCODE (*q* = 0.011), and coloc applied to SuSiE fine-mapped signals supported a shared causal variant in both cohorts (PP.H4=0.996 and 0.998). In both protein datasets, genetically higher SULT2A1 abundance was associated with a higher biliary-FU score. Cholelithiasis (K80), one of the 23 diagnoses encoded by this FU, also peaked within the same FU credible set in its UK Biobank GWAS.

Transcript and variant-level evidence added complementary support. Across 49 GTEx v8 tissues, the same MR and colocalization criteria supported 13 FU–gene pairs at ten loci in four FUs, including *CELSR2*, *FES* and *SORT1* from the plasma-protein set (Supplementary Fig. 1). Fine-mapped credible sets also contained three alleles with published functional evidence, the *IL33* splice-acceptor allele [18], the *FUT2* non-secretor stop allele [19] and rs12740374 at the *SORT1* locus [20]. Each lies in the genome-wide top 1% of predicted single-nucleotide variant impact in the annotation catalogue used here [21] (Supplementary Fig. 2). These annotations generate mechanistic hypotheses rather than assigning causal genes on their own.

Genes prioritized at FU loci are enriched for established therapeutic targets. Of 340 fine-mapped credible sets across eleven disease FUs, 272 contained a protein-coding gene with locus-to-gene score above 0.5, yielding 214 prioritized genes (Fig. 5c). Thirty of 214 were targets of approved drugs for any indication (odds ratio 3.29, 95% CI 2.15–4.90, *P* = 1.4 *×* 10*^−^*^7^), and 22 were approved for diagnoses encoded by these FUs (odds ratio 3.97, 2.41–6.24, Fig. 5d). Genes prioritized from the pooled ICD-10 diagnosis scans were also enriched (odds ratio 2.71 at the same 0.5 threshold), so the comparison supports therapeutic relevance of the FU gene set rather than superiority over diagnosis GWAS. Network propagation expanded 196 on-network L2G genes to 479 genes, including 68 approved targets, and the enrichment persisted after stratifying by network degree.

The cardiometabolic FU illustrates how the evidence layers connect (Fig. 5e). It encodes 83 diagnoses, 31 of which have MR-APSS support, and has 62 prioritized genes, eight of them approved drug targets (odds ratio 2.94, 95% CI 1.21–6.25). The displayed path includes established lipid targets such as *PCSK9*, *HMGCR* and *ANGPTL3*, while the broader prioritized set also includes *INSR* (L2G score 0.77), the receptor targeted by insulin in type 1 and type 2 diabetes. The atopic–immune airway FU provides a second example in Extended Data Fig. 8c. These examples show how shared disease structure can be carried from multimorbidity through genetic loci to molecular and therapeutic annotations without treating any single evidence layer as definitive.

## 2 Discussion

Starting from the multistage model of carcinogenesis and reliability theory, we asked whether the number of processes underlying a chronic disease could be measured independently from multimorbidity and used to explain variation in how incidence rises with age. The results support this idea. Across 232 diagnoses, diseases linked to more FUs showed faster incidence acceleration, and the relationship persisted when participant age was removed from the representation model. This turns the proposed disease complexity from a quantity inferred only from an incidence curve into an empirical property derived separately from disease co-occurrence. The same FUs carried distinct common-variant architectures whose directions were preserved in FinnGen and MVP, concentrated distributed genetic signal into regions missed by the specified diagnosis and BMI comparators, and connected through fine-mapping and molecular evidence to candidate genes enriched for approved drug targets.

The centrality–*r* relationship is the direct test of the motivating hypothesis. Multistage models explain differences in cancer incidence through the accumulation of multiple events, while reliability theory generalizes the principle to systems whose failure involves several components [1–3]. Here the proposed complexity is estimated from multimorbidity rather than read from the incidence curve. Among the 196 diagnoses with rising hazards, mean *r* increased from 2.50 at one linked FU to 6.09 at seven, and the same ordering was retained after removing participant age from the encoder and was stronger than for linear decompositions of the same diagnosis matrix. We interpret this ordering, rather than a fixed numerical slope, as the theoretical test. FU centrality remains a data-derived measure of how broadly a diagnosis engages the multimorbidity organization, not a literal count of microscopic failure events. Its biological relevance is supported by the learned basis itself: FU genetic correlations were lower than in random rotations, FU–diagnosis genetic correlations concentrated in the corresponding clinical chapters, and tissue and molecular profiles separated the dimensions. These observations support a reliability-like interpretation at the level of shared disease processes without requiring a one-to-one mapping between learned FUs and physical components.

Genetics provides both an independent test of this organization and a route from shared disease structure to mechanism. Each FU summarizes diagnoses that co-occur, so its GWAS emphasizes effects shared across those diagnoses while individual-diagnosis GWAS retain disease-specific signal. The external reconstructions show that this cross-disease genetic direction is preserved in FinnGen and MVP even when no matched UK Biobank diagnosis reaches genome-wide significance at the lead variant. The 19 disease-FU regions among the 34 comparator-unreached regions provide a complementary view: aggregating related diagnoses can raise weak but directionally coherent effects above genome-wide significance, as illustrated by the *KCTD16* -labelled locus. Earlier latent-phenotype studies established that joint disease representations can be heritable and yield additional associations [10, 12, 22]. Here those genetic properties are tied to an independently tested feature of disease incidence, and the molecular analyses provide a route from FU loci to testable gene-level hypotheses and established therapeutic biology.

Several features bound the interpretation. The FUs are learned from diagnoses recorded before UK Biobank assessment, and a zero in the matrix means no recorded diagnosis rather than confirmed absence of disease; healthcare contact, treatment and delayed ascertainment can therefore shape the representation. UK Biobank participation is selective [23, 24], and the reported FU heritabilities come from participating Europeans. The FUs themselves were not re-estimated in another cohort or ancestry group. External analyses instead test the direction of diagnosis-weighted genetic composites, supporting portability of the shared genetic pattern without constituting replication of the latent phenotype. Recruitment after midlife and first-record dates can also distort fitted age hazards, especially for diseases commonly present before assessment. These limitations motivate external relearning of the FUs, longitudinal representations that distinguish trajectories from accumulated burden, and tissue-resolved or experimental follow-up of prioritized loci.

Taken together, the results extend a principle familiar from multistage carcinogenesis to chronic disease: variation in age-dependent incidence can be related to disease complexity measured independently from multimorbidity rather than inferred from incidence alone. The resulting functional organization lies between individual diagnoses and global measures of ageing. FU centrality describes how broadly each diagnosis is embedded in that organization, while genetics shows that the same dimensions carry distinct and portable biological architectures and can concentrate shared signal across conventional disease boundaries. The FUs should therefore be viewed neither as diagnostic categories nor as literal microscopic failure events, but as empirically defined dimensions through which multimorbidity, age-dependent incidence and shared genetic biology can be studied on a common disease scale.

## 3 Methods

### 3.1 UK Biobank cohort and ethics

UK Biobank recruited approximately 500,000 participants in 2006–2010 with electronic signed consent at 22 assessment centres in England, Scotland and Wales [9]. The modelling cohort comprised 502,355 participants, irrespective of genetic ancestry or number of diagnoses. Participant-level genetic analyses of the FUs used the 423,955 participants of the European genetic-ancestry group after quality control, related participants included, unless stated otherwise.

A three-character ICD-10 code counted as chronic when the Chronic Condition Indicator Refined for ICD-10-CM v2026.1 [25] flagged any of its ICD-10-CM codes as chronic, and codes without an ICD-10-CM counterpart in the indicator were classified by hand. The disease panel comprised the 232 such codes that were not cancer or near-cancer codes, had a prevalence at assessment of at least 0.001 and had a male fraction among cases between 0.10 and 0.90 inclusive.

### 3.2 Electronic health record data sources and preprocessing

Diagnosis ages came from the UK Biobank first-occurrence fields, which combine linked primary-care, hospital inpatient and death-register records with self-report at assessment [26], and cancer diagnoses from the cancer registry. With *a_i_* the age at assessment and *t_ij_* the age at first record of diagnosis *j*, the model input was

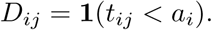

A zero therefore denotes no diagnosis recorded before assessment, not established absence of disease.

### 3.3 Age-specific incidence and hazard fitting

For each diagnosis, follow-up ran from assessment to its first record, death or 1 January 2016, whichever came first, because primary-care records end in 2016–2017. Incidence in each three-year age band from 44–46 to 74–76 was the number of first diagnoses divided by the person-years at risk in that band. Cancer-registry diagnoses C00–C97 other than C44 were grouped by ICD-10 malignant-neoplasm block, with the first diagnosis in a group as its event and the same follow-up. The nine groups drawn in Fig. 2a fall within the sex-fraction band of the disease panel and were recorded in at least 0.1% of participants, with secondary and ill-defined sites excluded, and serve only as reference curves. For each diagnosis the cause-specific hazard

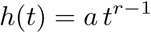

in attained age *t* was fitted by maximum likelihood with left truncation at assessment, with *a >* 0 and *r* unrestricted in sign. For *r >* 0 this is the Weibull hazard with shape *r*. Intervals are 95% profile-likelihood intervals. The centrality analysis used two diagnosis sets defined by the hazard fit alone, all 232 diagnoses and the 196 with *r̂ ≥* 1, the range to which reliability theory applies. The association is read as consistent with that theory, not as evidence for it. Survivor selection at recruitment and the dating of diagnoses at first record can bias the association of *r̂* with FU centrality in a direction that this cohort cannot bound.

FU centrality, the number of FUs linked to a diagnosis (§3.6), depends on the *L*_1_ penalty, which was chosen within the analysis. Among grid penalties giving a mean centrality between 1 and 15 over the 232 diagnoses, a criterion free of *r*, the one with the largest Spearman correlation between centrality and *r̂* was selected in each set, ties going to the larger penalty. The *P* value carries this choice. It is the share of 10^5^ permutations of centrality within ICD-10 chapters, each moving a diagnosis’s centralities at all candidate penalties together, in which the largest absolute correlation over all candidate penalties, each centred on its permutation mean, reached the observed correlation at the selected penalty centred in the same way, so it concerns association within chapters, and each set was tested at 0.025. Confidence intervals came from a within-chapter bootstrap of diagnoses at the selected penalty and are conditional on it, with the autoencoder and the hazard fits held fixed. Selection on random within-chapter halves of the diagnoses, evaluated on the other halves, gauged optimism and is not a replication. The test was repeated on ICD-10 block means and with one chapter left out at a time. The rule selected *λ* = 0.0794 on both sets, with 804 FU–diagnosis pairs and a mean centrality of 3.47. The diagnosis sets, the rule, the test levels and the follow-up end date were fixed after correlations with centrality had been seen on earlier hazard fits. Spearman’s *ρ* was 0.472 among the 183 diagnoses whose interval for *r* lay above 1, 0.463 among the 205 whose interval did not lie below 1, 0.485 among the 216 with *r̂ >* 0 and 0.562 among the 165 with at least 1,000 incident cases, the last at its own selected penalty. When follow-up instead ended at the end of hospital records in each country, *ρ* was 0.515 over all 232 diagnoses and 0.458 over the 183 diagnoses with *r̂ ≥* 1 in those refitted hazards.

As a sensitivity analysis, an autoencoder with the same architecture and training settings was trained without age at assessment as an input, and 15 of its latent dimensions passed the activity threshold of §3.4. Their centrality, computed as for the FUs with candidate mean counts between 1 and 14, was correlated with *r̂* at *λ* = 0.0794, with the path-adjusted permutation *P* value and bootstrap intervals that do not include training uncertainty.

### 3.4 Latent phenotype modelling

A variational autoencoder was trained on 232 binary diagnosis indicators and body mass index (BMI). The encoder also received a feature-observation mask, age at assessment and sex, and the decoder the latent vector and sex. Age entered only the encoder, so FU scores are age-informed by construction. Age at first diagnosis and genotype were not inputs. The approximate posterior was a 32-dimensional diagonal Gaussian, and training maximized the evidence lower bound

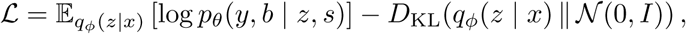

with *y* the diagnoses, *b* BMI, *s* sex and *x* all encoder inputs. Diagnoses had Bernoulli likelihoods and BMI a Gaussian likelihood with one learned variance shared by all participants. BMI was standardized and clipped at its observed 1st and 99th percentiles. Reconstruction terms were summed over observed features and averaged over participants, and the Kullback–Leibler term had unit weight. A coordinate was retained as an FU when its mean marginal Kullback–Leibler divergence from the prior exceeded 0.05, which left 16 of 32. Hyperparameters were chosen by validation loss on a random 20% of participants (Extended Data Fig. 1c,d). Downstream analyses used the posterior mean of each FU, except Extended Data Fig. 2a, which used one sampled posterior value. The fitted model served as a fixed descriptive representation, not as a causal or clinical-prediction model.

### 3.5 Ten-year FU displacement and participant map

A participant’s displacement is the difference between the encoder’s scores for their diagnosis record at assessment and for the same record ten years later, over the 15 disease FUs. BMI is unavailable at follow-up, so numerical summaries used baseline scores re-encoded with BMI masked. They cover the 237,704 participants aged at most 69.8 years at the follow-up point whose follow-up scores agreed with those from the reconstructed encoder, and Fig. 2e shows the 147,839 of them with at least one new diagnosis. Fig. 2d pools the assessment visit and later two-yearly timepoints of each participant, with BMI carried forward from assessment and scores standardized on all participants at assessment, and gives the fraction of records above +1 s.d. at each age.

The participant map is a uniform manifold approximation and projection (UMAP) of the 16 standardized FU scores of the 293,061 participants with at least two diagnoses. Extended Data Fig. 1a,b colours lattice cells holding at least 30 participants by a cell mean of a baseline quantity, and Fig. 2c by the most common dominant ICD-10 chapter, the chapter holding most of a participant’s diagnoses. Streamlines are ten-year displacements of a fixed sample of 80,000 mapped participants, projected into the map, averaged within cells and smoothed. Unlike the numerical summaries, they compare baseline scores computed with BMI with ten-year scores computed without it, so part of each streamline reflects the missing BMI input rather than new diagnoses. A control with structureless random displacements reproduced most of the projected field, so its visual organization largely reflects the neighbour graph and the projection geometry. The map is built from the diagnoses that define the FUs and describes the representation.

### 3.6 FU definition and phenotypic characterization

“Functional unit” denotes a learned collective dimension of the diagnosis matrix. The coordinates were learned without expert-defined clinical groupings. Descriptive names were assigned after fitting from each FU’s leading diagnoses and ICD-10 chapter associations, with genetic, biomarker and medication results consulted, and a mechanism word such as autoimmune was kept only where the FU’s GWAS loci also carried it. Each FU was oriented by its mortality association so that higher values mean worse survival.

Chapter associations (Fig. 2b) came from logistic regressions of chapter case status on one FU at a time, adjusted for age, sex, smoking status and Townsend deprivation index, with Benjamini–Hochberg correction over the 192 FU–chapter tests.

For each of the 16 *×* 232 FU–diagnosis pairs, an *L*_1_-penalized logistic regression of diagnosis status at assessment on one FU score, age and sex, all standardized, was fitted in all 502,355 participants. Every coefficient, the intercept included, was penalized, and the objective was the mean class-weighted negative log-likelihood, with balanced class weights, plus *λ* times the *L*_1_ norm of the coefficients. The FU score used for selection was target-masked, computed by the trained encoder with the target diagnosis set to its median, zero, and marked unobserved in the encoder mask. The encoder was trained on the same participants, so masking does not make selection out-of-sample. At the selected penalty (§3.3) 804 pairs had a non-zero FU coefficient. A diagnosis linked to, retained by or encoded by an FU belongs to one of these pairs, and FU centrality counts them.

Blood count, clinical chemistry and Olink plasma proteins measured at assessment characterized the FUs and did not define them. Each standardized analyte was regressed on one FU at a time by least squares with robust standard errors, adjusted for age, sex, ethnicity, Townsend deprivation index, smoking status, assessment centre, fasting time and season. Benjamini–Hochberg correction and Bonferroni flags used each assay grid, and filled points of Extended Data Fig. 3a also exceed a minimum absolute coefficient. These are marginal cross-sectional associations.

For medication sensitivity, analyte models were refitted on identical participants with and without ten self-reported medication-class flags, and the change in the FU coefficient was decomposed over the flags by the Gelbach identity. A displayed association carries a grey class label when one class accounts for more than half of the FU coefficient and has an established effect on the analyte in the direction of the class’s own coefficient. The shares are point estimates without confidence intervals, and a flag marks the indication as well as the drug. Medication use in the 10% of participants at the disease end of each FU, the end where its named diagnoses are more common, was compared with the other participants by an observed-to-expected ratio standardized for sex and age, with Cochran–Mantel–Haenszel tests and Benjamini–Hochberg correction over the 160 FU–class cells.

### 3.7 All-cause mortality model

All 502,355 participants were followed from assessment to death or 12 November 2021. Cox models used time since assessment as the timescale with age at assessment and sex as covariates, and the 15 disease FUs were standardized and entered jointly. Proportional hazards were not assessed. Hazard ratios per standard deviation come from this joint model fitted in all participants, with Wald intervals. The FU risk score is the linear predictor of the FU terms. Its weights and all compared models were estimated by five-fold cross-validation, and pooled held-out predictions gave one Harrell’s *C* per model, with intervals for differences from a paired bootstrap of these predictions. Models contained age and sex alone or with the prevalent-diagnosis count, the FU risk score, both, or the 232 diagnosis indicators with and without the FU risk score. The autoencoder was not refitted within folds.

### 3.8 Linear representation comparators

Logistic principal-components analysis and logistic singular-value decomposition, each with 20 components and age- and sex-based offsets, were fitted to the same diagnosis matrix. Component scores were target-masked, exactly for logistic PCA and approximately for logistic SVD, and component centrality was analysed with the per-pair *L*_1_ models, selection rule, permutation, bootstrap and split-half procedures of the FU analysis, with candidate penalties at a mean count between 1 and 19. Components were fitted on one subset of participants and component counts estimated on a separate association split of 100,471, whereas FU centrality used all 502,355. On the association split the FU rule selected the same penalty as in all participants, which is not a hold-out because the autoencoder was trained and validated on all participants (Extended Data Fig. 2c,d). The FU–component correlations of Extended Data Fig. 2a are Spearman correlations, and its joint *R*^2^ comes from regressing FU midranks on the midranks of all 20 components with an intercept. They use components fitted on all 502,355 participants and are descriptive.

### 3.9 FU genetic correlations

Observed-scale SNP heritability and genetic correlations were estimated by LD score regression [27] with a free intercept, on HapMap3 variants with Pan-UK Biobank European LD scores [15], for the 120 distinct FU pairs and for FU–diagnosis pairs with this study’s 232 diagnosis scans. Benjamini–Hochberg and Bonferroni correction ran over the 120 FU pairs and, separately, over all estimable FU–diagnosis cells. A chapter named by an FU counted as concentrated when it held strictly more false-discovery-rate-significant FU–diagnosis correlations than any other ICD-10 code-letter chapter, ties counting as not concentrated. For a descriptive comparison, the genetic covariance matrix *G* = *D*^1^*^/^*^2^*R_g_D*^1^*^/^*^2^, with standalone heritabilities in *D* and bivariate genetic correlations in *R_g_*, was expressed in 2,000 Haar-uniform random orthonormal bases and restandardized to correlations after each rotation, which gives no calibrated *P* value. These correlations reuse the participants and diagnoses that define the FUs and do not validate the FU labels independently.

### 3.10 FU–diagnosis Mendelian randomization

Directional genetic coherence between FUs and their linked diagnoses was estimated with MR-APSS [17], which models sample overlap and correlated pleiotropy through a background component from LD score regression. Exposure and outcome GWAS came from the same 423,955 European-ancestry participants. The family was the 804 linked pairs, including 40 BMI-FU positive-control pairs. Instruments were clumped FU variants at *P ≤* 5 *×* 10*^−^*^5^ with selection-bias correction, and Benjamini–Hochberg correction ran over the 804 tests. MR-Egger and weighted-mode estimates, on at least three clumped instruments at *P <* 5 *×* 10*^−^*^8^ and available for 587 pairs, described direction and their own significance, with Benjamini–Hochberg and Bonferroni correction over each estimator’s 587 tests. Unlike MR-APSS, these two estimators did not model sample overlap. MR-APSS was also run on the 40 BMI-FU diagnoses with the Pan-UK Biobank BMI scan [15] and the GIANT–UK Biobank BMI meta-analysis [16] as exposures, with the same settings and Benjamini–Hochberg correction over the 40 tests of each exposure (Extended Data Fig. 9). Pair selection used target-masked scores and the exposure GWAS unmasked posterior means. The outcome diagnoses contribute to FU construction, so the estimates test genetic coherence under the MR-APSS assumptions and are not evidence that an FU causes its constituent diagnoses.

### 3.11 Genotyping and ancestry assignment

Participants were excluded on the sample quality-control flags of the central UK Biobank genotyping workflow [9]. Six genetic-ancestry analysis groups came from the Pan-UK Biobank labels [15], European (423,955), Central/South Asian (8,952), African (6,698), East Asian (2,680), Middle Eastern (1,533) and Admixed American (922). These are genetic-analysis groupings, distinct from self-reported categories.

### 3.12 Genome-wide association analyses of FUs

REGENIE v4.1 [28] tested the posterior mean of each of the 16 FUs as a quantitative trait in the 423,955 European-ancestry participants, related participants included, with quality-controlled, LD-pruned genotyped variants in step 1 and imputed variants passing imputation-quality and allele-frequency filters in step 2. Covariates were sex, age, age squared, their interactions with sex, the first 20 genetic principal components of UK Biobank Field 22009, genotyping array and assessment centre. The 232 diagnosis scans used REGENIE binary-trait tests with approximate Firth correction and the same covariates except genotyping array. Genome-wide significance was *P <* 5 *×* 10*^−^*^8^. Loci were runs of significant variants with minor allele frequency at least 0.01 separated by less than 500 kb, each led by its most significant variant, 979 over the 16 scans.

A model with the same sex and age terms and 20 genetic principal components, without assessment centre or genotyping array, was run in the five smaller ancestry groups. These were compared with the European results by direction of effect at the FU leads of the 1,102 regions of §3.14 and at frequency-matched control variants at least 1 Mb from any genome-wide-significant variant of the same FU (Extended Data Fig. 6a,b). These groups are not powered for genome-wide replication, and no discovery or downstream result depends on them.

### 3.13 External direction test in FinnGen, MVP and CKB

The test asks whether a composite of an external cohort’s own diagnosis scans keeps the direction of the UK Biobank FU association at FU lead variants. It used published summary statistics from FinnGen R12 [29], the Million Veteran Program (MVP) release 4 European analysis [30] and China Kadoorie Biobank (CKB) [31], with high-confidence matches of UK Biobank diagnoses to endpoints of at least 500 cases. The eleven FUs tested are those other than the BMI FU with a lead outside the MHC at *P <* 1 *×* 10*^−^*^8^. Composite weights came from a ridge regression through the origin of the UK Biobank FU *Z* on the UK Biobank *Z* of the matched diagnoses at LD-pruned HapMap3 variants, with the chromosome of each lead held out, and were applied to the cohort’s endpoint *Z* at the lead. For each FU and held-out chromosome, the ridge penalty, with an unpenalized fit among the candidates, was chosen by five-fold cross-validation over groups of the remaining chromosomes, minimizing prediction mean-squared error averaged equally over validation chromosomes, and predictors were scaled by their training-set root-mean-square without centring. Each composite was scaled by its root-mean-square over background variants, five per lead–FU nomination, matched on minor-allele-frequency decile and at least 1 Mb from any lead and any genome-wide-significant variant of the same FU. In FinnGen a lead missing from the release was replaced by a proxy in strong linkage disequilibrium, in MVP a missing endpoint contributed zero, and where MVP reported *P* = 0, *Z* was derived from the effect estimate and its confidence interval. Leads that could not be scored, leads removed to keep tested leads independent (*r*^2^ *<* 0.01 within each FU in FinnGen and MVP, 1 Mb across FUs in CKB) and leads whose missing endpoints carried more than 10% of the weight or the top weight were not tested, which left 257, 257 and 181 of 321 lead–FU nominations in FinnGen, MVP and CKB. A lead passed when its two-sided *P* was below 0.05 divided by the number of tested leads. One-sided exact sign-flip tests over chromosomes, each chromosome counted once, one-sided exact binomial tests per FU and one-sided two-sample Fisher tests of pooled agreement against background agreement summarize the leads (Fig. 3f,g, Extended Data Fig. 6c–f). The composite is not a measurement of the FU in the external cohort, and the test concerns direction only. The genome-wide tracks of Extended Data Fig. 4b are descriptive. Their marked leads were matched on position and alleles without the insertion– deletion identity check of the lead test, so they include untested nominations, and a missing endpoint or one with an odds ratio of 1 counts as zero and shrinks a variant’s scale, which can raise single MVP peaks. CKB was exploratory because of its smaller endpoint set and different ancestry, and its autoimmune-thyroid composite does not represent that FU because no CKB thyroid endpoint reaches 500 cases.

### 3.14 Genomic-region recovery

FU and comparator scans were compared on one shared partition, with region centres at least 500 kb apart chosen greedily from variants at *P <* 5 *×* 10*^−^*^8^ and minor allele frequency at least 0.01 pooled across the FU scans, the comparator scans and Pan-UK Biobank scans screened for the comparators but not admitted to them. A scan reached a region when it had a qualifying variant within 500 kb of the centre, so one locus can reach several regions. Of 1,716 regions the FU scans reached 1,102. Removing every region reached by this study’s 232 diagnosis scans, the Pan-UK Biobank BMI scan, the GIANT–UK Biobank BMI meta-analysis [16] or 228 Pan-UK Biobank scans matched to the 232 codes directly or through one documented phecode proxy left 34. Half-widths of 750 and 1,000 kb were sensitivity analyses. The 34 regions are a set difference relative to these comparators, several of which draw on UK Biobank. Membership in the 34 is specific to the diagnosis codings scanned, and broader published definitions of asthma and knee osteoarthritis reach genome-wide significance at *RIN3* and *USP8*, respectively [32, 33]. The 34 were not tested for enrichment as a set, the mechanism that brings them to significance was not isolated, and they are neither an independent replication nor an estimate of net locus gain. Region names are nearest protein-coding genes, a positional label. Seven of the 34 regions carry a scored FU credible set. At 22 others the lead lies between 1 *×* 10*^−^*^8^ and 5 *×* 10*^−^*^8^, above the fine-mapping threshold of §3.19, and at five fine-mapping returned no credible set in the region.

At the *KCTD16* region the associations of the 83 diagnoses that the cardiometabolic FU encodes were compared at the FU lead, to ask whether the FU signal reflects many weak concordant diagnosis effects (Fig. 4c). The diagnoses share cases, so their associations at one variant are correlated, and counts of concordant diagnoses are descriptive. External composites at the region leads were descriptive. GWAS-catalogue records were collected at the nine region leads of the cardiometabolic and atopic– immune airway FUs and at their proxies with *r*^2^ *≥* 0.8 within 500 kb, from the GWAS Catalog release of 22 August 2026 and GWAS Atlas, counting one association per lead, phenotype and catalogue and a proxy’s phenotype once per lead, only when the lead carried it in neither catalogue.

### 3.15 Gene and pathway scoring

Gene-level *P* values were computed from each FU scan with PascalX [34], which aggregates variant associations around each gene while accounting for linkage disequilibrium, with Benjamini–Hochberg correction over genes within each FU. Pathways from eight MSigDB collection groups [35, 36] were scored from the gene results with the PascalX chi2rank statistic, with Benjamini–Hochberg correction within each FU and collection. The MHC was kept in pathway scoring. Because its linkage disequilibrium can count one signal more than once, pathways were also rescored without MHC genes in the sets and the ranking. Odds ratios beside pathways counted set members within 100 kb of one another as one unit, significant when any member had a gene-level false-discovery rate below 0.05, against the significant and other tested genes of the FU outside the set, with approximate Woolf intervals. They describe the density of gene-level signal and are not tests. Redundant Gene Ontology terms were reduced by semantic similarity [37] for display only. Collections overlap, so agreement between them is not independent evidence.

### 3.16 Tissue specificity

For each FU and each of 49 GTEx v8 tissues [38], gene-level *Z* scores, Φ*^−^*^1^(1 *− P*), were regressed on the tissue’s expression over scored genes outside the MHC. Median expression was capped at 50 transcripts per million, transformed as log_2_(1 + TPM) and standardized across genes within each tissue. Covariates were log gene length, log number of tested variants within 50 kb, log of one plus the number of neighbouring genes within 1 Mb and mean transformed expression across tissues, and both expression and gene *Z* were residualized on them with an intercept. One-sided *P* values came from circular shifts of the residualized expression along the genes in genomic order, which preserve the correlation between neighbouring gene scores. Bonferroni and Benjamini–Hochberg thresholds were applied over the 784 FU–tissue tests. Bulk expression resolves tissues rather than cell types.

### 3.17 Cis molecular evidence and colocalization

*Cis*-Mendelian randomization and colocalization tested the assayed proteins of the UK Biobank Pharma Proteomics Project (Olink) [39] and deCODE (SomaScan) [40] panels against the scans of the 15 FUs other than the BMI FU, one assay measurement at a time, so a supported gene need not lie at a fine-mapped FU locus. The primary estimator was MR-Link-2 [41], which uses the whole *cis* region and models horizontal pleiotropy and linkage disequilibrium, and support required Benjamini– Hochberg *q <* 0.05 within each FU and cohort over the whole assay panel. Inverse-variance-weighted MR on clumped *cis* instruments at *P ≤* 5 *×* 10*^−^*^8^, corrected within each FU and cohort over the reagents with an instrument, was a comparator that selects no pair of Fig. 5a. Colocalization used SuSiE [42] where both traits returned a credible set and the approximate Bayes factor [43] otherwise, never the larger of the two, with a fixed minor-allele frequency of 0.5, the median sample size of each trait, each trait’s standard deviation estimated from its effect variances under these two values and priors *p*_1_ = *p*_2_ = 10*^−^*^4^ and *p*_12_ = 10*^−^*^5^ for the approximate Bayes factor. Colocalization support required a shared-variant posterior of at least 0.8. A pair in Fig. 5a meets both supports in the same assay measurement. The plasma board adds pairs that colocalize in the same assay and are supported by inverse-variance-weighted MR on more than one instrument, and Supplementary Fig. 1 draws the GTEx readout of its pairs. UK Biobank proteomics shares participants with the FU scans, whereas deCODE is independent. For proteins measured on both panels, the lead *cis* instrument was looked up in the other panel and counted as replicated when it had the same direction at *P <* 5 *×* 10*^−^*^8^ there. Plasma-protein evidence cannot separate effects on abundance from effects on function or assay binding.

The same estimators were applied to GTEx v8 *cis*-eQTLs in 49 tissues from the eQTL Catalogue [38, 44], for genes with a *cis* association at *P ≤* 5 *×* 10*^−^*^5^ and with Benjamini–Hochberg correction over the genes with an estimate within each FU and tissue, separately for each estimator (Supplementary Fig. 1), and to whole-blood eQTLGen [45]. Supported GTEx pairs need not lie at genome-wide-significant FU loci. Counting every gene submitted to MR-Link-2 instead reduced the supported set from 16 to 13 FU–gene–tissue combinations and from four to three FUs. Transcript results are reported beside the protein results and select no pair of Fig. 5a.

### 3.18 Network-based signal propagation

Locus-to-gene scores were propagated by a random walk with restart, restart probability 0.4, through the largest connected component of a high-confidence Open Targets protein-interaction network. Seeds were each gene’s maximum locus-to-gene score in the FU, kept as weights only at scores of at least 0.5. A non-seed gene was added when its score ranked in the top 1% for the FU and its one-sided empirical *P* over 10,000 permutations of the seed weights was below 0.01. Propagation was carried forward for the six FUs with more than ten seed genes in the network, the BMI FU and five disease FUs. Added genes are network-prioritized groups without error-rate control for individual genes. With restart probabilities from 0.01 to 0.95 and seed thresholds of 0.5 or 0.8, the seed and propagated genes remained enriched for approved targets, all 12 degree-stratified odds ratios having 95% intervals that exclude 1. With every locus-to-gene score used as a seed weight and no cutoff, the seed and propagated genes were not enriched for approved targets.

### 3.19 Target prioritization and therapeutic enrichment

Leads and regions used *P <* 5 *×* 10*^−^*^8^, and fine-mapping and locus-to-gene scoring used signals at *P <* 1 *×* 10*^−^*^8^ outside the MHC. Credible sets came from SuSiE-inf with Pan-UK Biobank European linkage disequilibrium, and genes were scored with the Open Targets locus-to-gene model [46, 47], whose local implementation, rerun on 25 GWAS Catalog studies that Open Targets has ingested, recovered 98.6% of the genes that the published Open Targets outputs call at a score of at least 0.5. A gene was prioritized when its score exceeded 0.5, with 0.8 and 19 cutoffs from 0.05 to 0.95 as sensitivity analyses. The 0.5 cutoff is an operating point and is not calibrated for the locus-to-gene model release used here.

A gene was an approved target when a ChEMBL 36 [48] phase-4 drug–indication record mapped to it through the drug’s mechanism. Enrichment was tested by two-sided Fisher exact tests against 20,125 protein-coding genes for approval in any indication, for a diagnosis encoded by the eleven disease FUs with scored genes and for a diagnosis among the MR-APSS-supported links. Diagnoses were matched to indications through an ICD-10–ontology crosswalk and genes were pooled across FUs, so a gene need not have been prioritized by the FU that encodes the matching diagnosis. *P* values are nominal. The rows of Fig. 5d are nested and were not compared by test, and the pooled diagnosis scans and the BMI FU are comparators, not superiority tests. The pooled diagnosis-scan comparator comprised the 72 diagnosis scans that yielded a credible set. Network sets were tested on a protein-coding network background without the top 1% of genes by degree, with Mantel–Haenszel odds ratios stratified by degree quintile. Fig. 5e illustrates the cardiometabolic FU with a subset of its MR-APSS-supported encoded diagnoses and genes chosen by explicit display rules. A gene is admitted when MR-Link-2 or colocalization supports it, or when it is the target of a drug approved for a diagnosis the FU encodes, so for a gene admitted through a drug the drug is the selection criterion and not a result. The rule was applied to the 20 genes examined for this FU, not to all 127 of its prioritized and propagated genes, and in Extended Data Fig. 8c to 25 of the 120 genes of the atopic–immune airway FU.

### 3.20 Variant-impact annotation

Credible-set members of the eleven disease FUs were annotated with the AlphaGenome Atlas variant-impact PHRED score [21] by exact GRCh38 position and allele. The score ranks all possible single-nucleotide variants, with 20 marking approximately the top 1%. Insertions and deletions are unscored. The score assigns no gene, tissue, direction or causal probability and did not alter fine-mapping posteriors (Supplementary Fig. 2).

### 3.21 Clinical-stage annotation

ChEMBL phase-4 labels annotate overlap of prioritized genes with targets of approved drugs, and Fig. 5e marks targets of drugs in trials with their highest trial phase (ChEMBL through Open Targets). Neither is an analysis of clinical-phase transitions or drug-development success.

### 3.22 Statistical analyses

Multiple-testing correction was applied within the analysis families stated above, not across the manuscript. Tests were two-sided unless described above as one-sided.

## Extended Data

**Extended Data Fig. 1.**
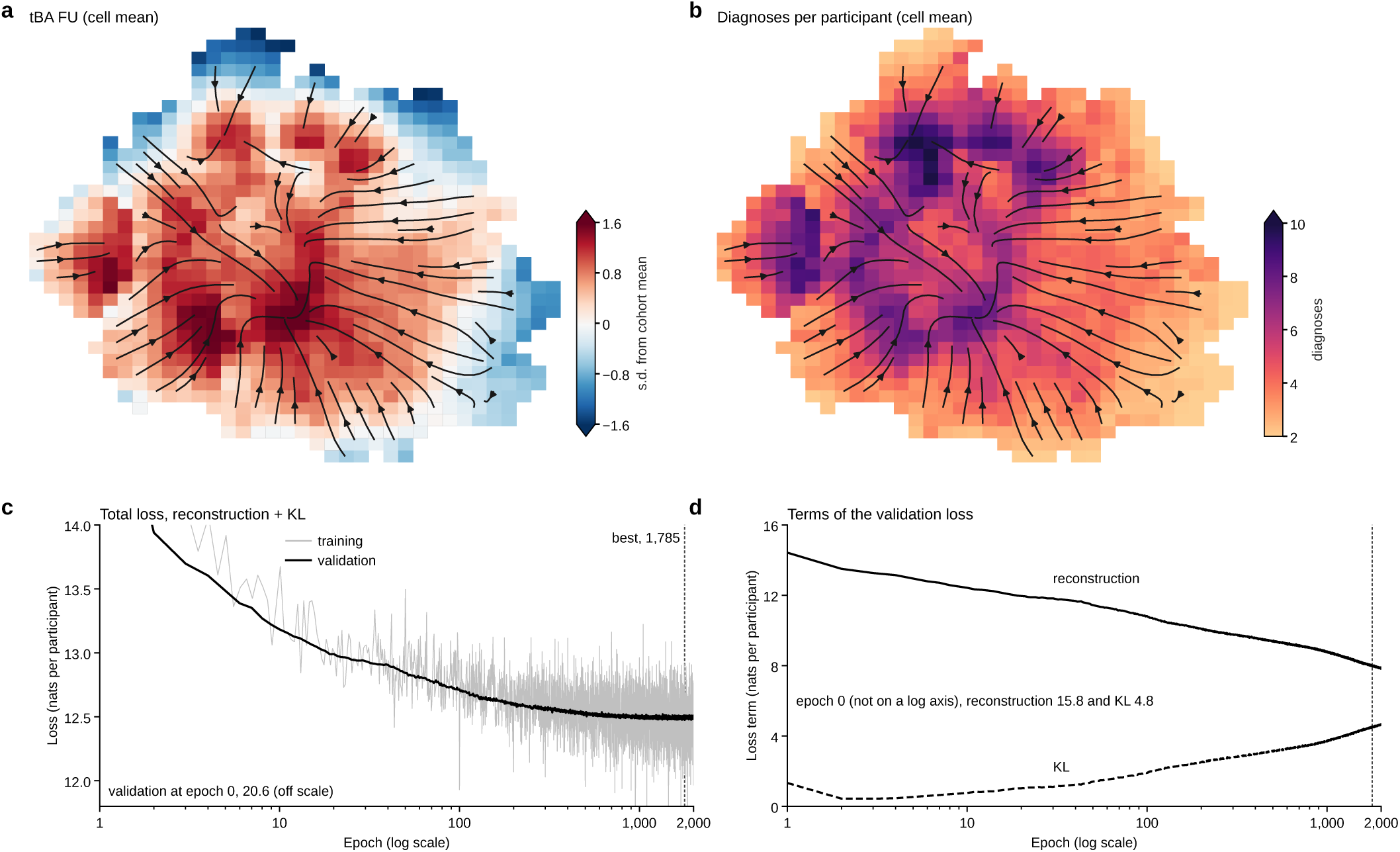
Thermodynamic biological age (tBA) broadly tracks disease burden on the participant map while retaining distinct structure. Panels a and b use the same UMAP of 293,061 participants with at least two diagnoses and colour a 36 *×* 36 lattice by baseline cell means. The layout is for visualization only and its axes have no units. streamlines show projected average ten-year displacements of 80,000 mapped participants and are illustrative rather than participant trajectories. **a**, Mean tBA FU score, standardized to all 502,355 scored participants. **b**, Mean number of prevalent diagnoses. Across 837 populated cells the two measures correlate at *r* = 0.754, but their maxima occur in different locations. Methods give the UMAP, lattice and displacement construction. **c**, Variational autoencoder total loss by epoch. Validation used a random 20% of participants (100,471), not a held-out test set. Training shows single mini-batches every 50 steps. The vertical line marks the best epoch, 1,785. **d**, Validation reconstruction and Kullback–Leibler (KL) terms.

**Extended Data Fig. 2.**
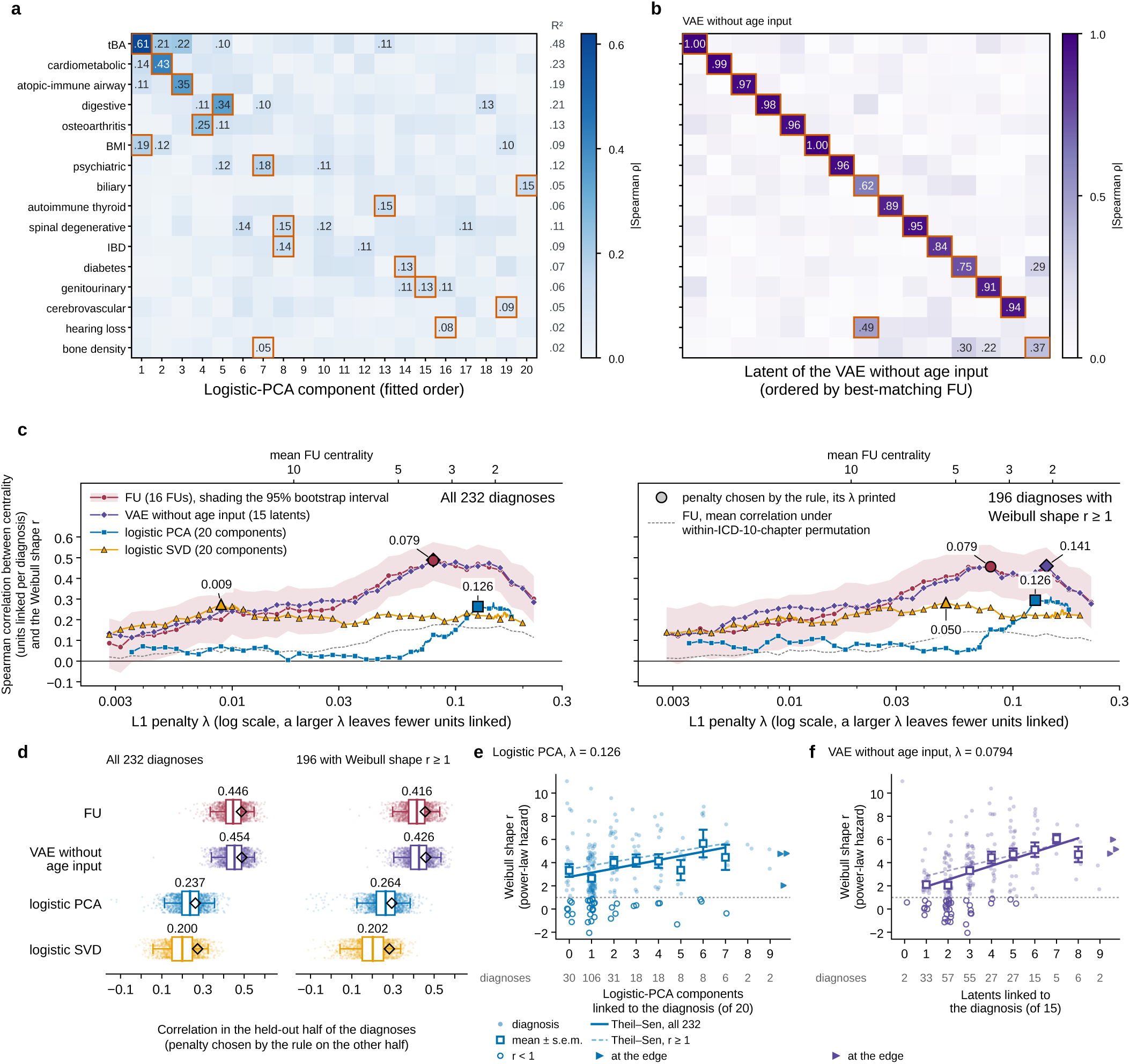
The centrality–incidence relationship persists without age input and is weaker for linear diagnosis decompositions. **a**, Absolute correlation between sampled FU values and 20 logistic-PCA components in all 502,355 participants. Outlines mark each FU’s strongest component match and the right column gives the joint *R*^2^ of all 20 components. b, Absolute correlation between primary FU posterior means and the 15 active dimensions of the VAE retrained without age. outlines mark the strongest match for each FU. **c**, Spearman *ρ* between component count and age-scaling parameter *r* across the *L*_1_ penalty path for the primary VAE, no-age VAE, logistic PCA and logistic SVD on the 100,471-participant association split. The FU penalty was chosen on all 502,355 participants and both VAEs were trained on all participants, so the split is not a hold-out. **d**, Split-half penalty transfer. Each point is the correlation in one diagnosis half at the penalty selected the in other. Boxes show interquartile ranges and whiskers the 2.5th–97.5th percentiles. **e**, Age-scaling parameter *r* against retained logistic-PCA components at its selected penalty. **f**, Age-scaling parameter *r* against no-age VAE centrality in all 502,355 parrticipants at the fixed primary penalty. Squares are level means *±* s.e.m. and lines are Theil–Sen fits.

**Extended Data Fig. 3.**
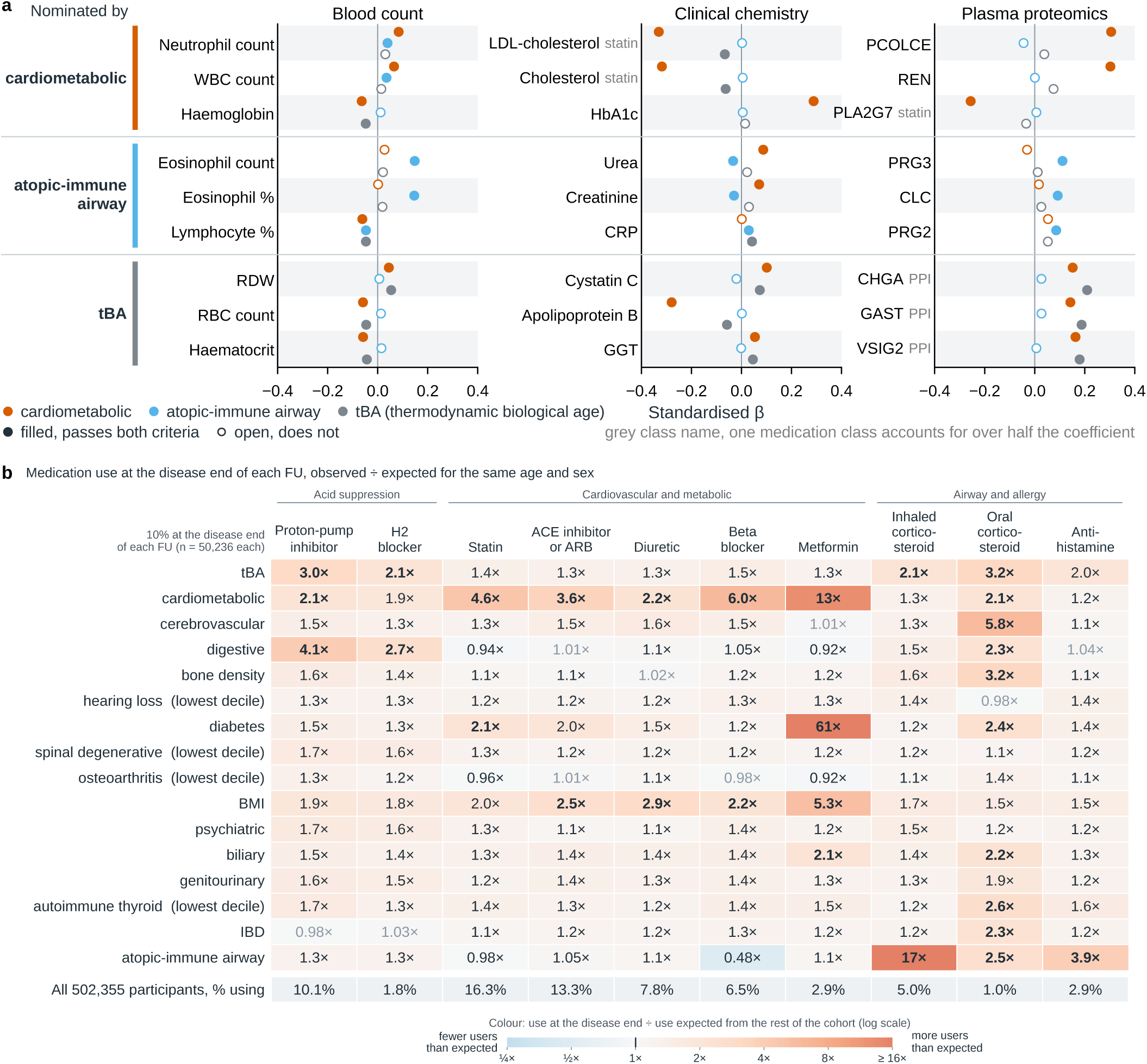
Selected functional units (FUs) have distinct molecular profiles, with treatment contributing to part of the cross-sectional signal. **a**, Associations of the cardiometabolic, atopic–immune airway and tBA FUs with selected blood-count, clinical-chemistry and Olink protein measurements. Points are covariate-adjusted effects per s.d. of the FU with 95% CIs. Filled points pass the assay-wide Bonferroni threshold and a minimum-effect filter. Grey medication labels identify displayed associations for which one drug class accounts for more than half of the FU coefficient in the medication decomposition described in the Methods and has an established effect on the analyte in that direction. **b**, Use of ten medication classes in the 10% of participants at the disease end of each FU, relative to the other 90% after age and sex standardization. colour is the observed-to-expected ratio on a log_2_ scale. Bold cells have 95% intervals wholly outside 0.5*×* to 2*×*, and light grey cells do not pass 5% FDR. These ratios characterize treatment patterns associated with the recorded diagnoses and are not estimates of drug effects. ACE, angiotensin-converting enzyme. ARB, angiotensin II receptor blocker. IBD, inflammatory bowel disease. PPI, proton-pump inhibitor. tBA, thermodynamic biological age.

**Extended Data Fig. 4.**
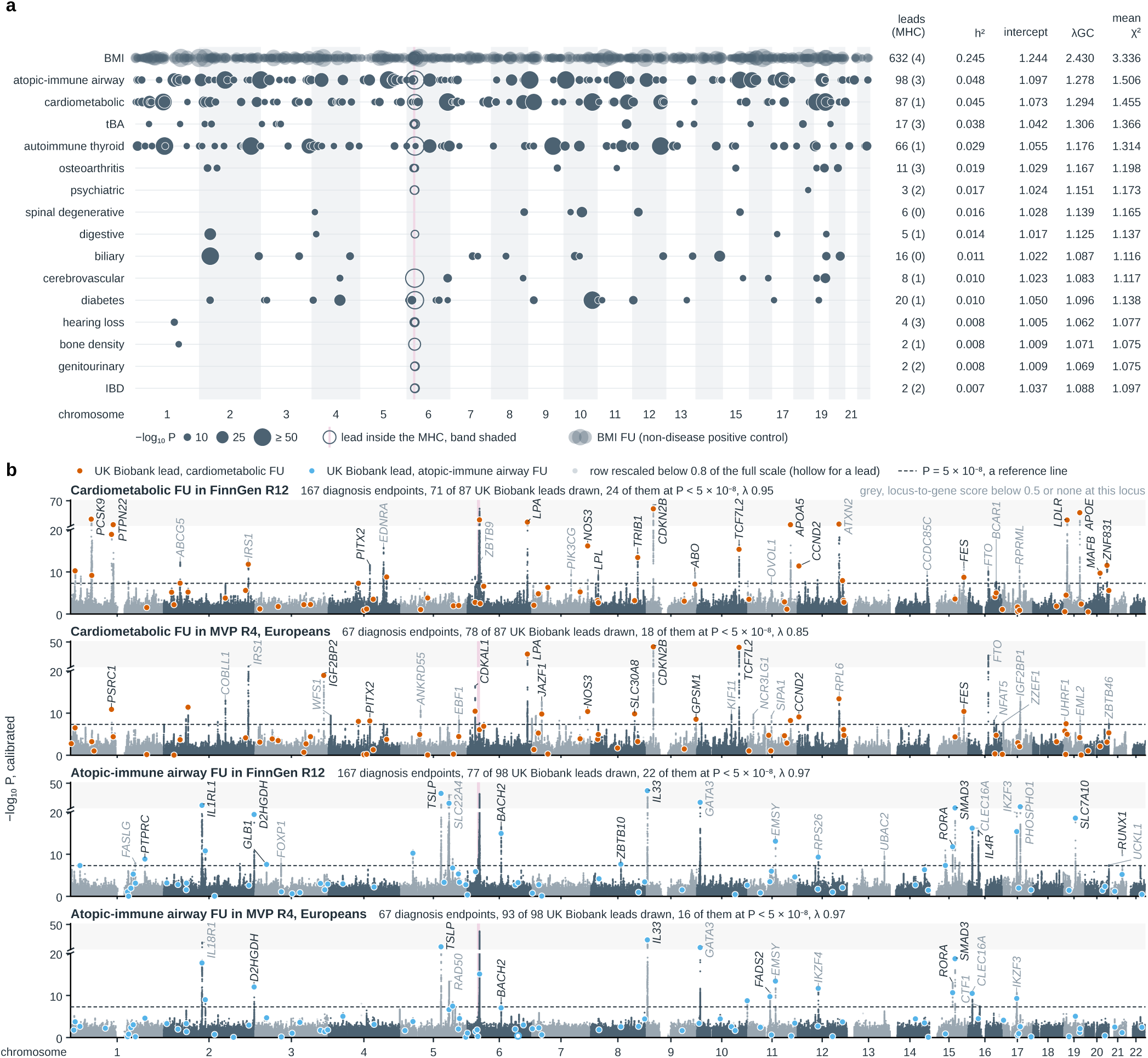
The 16 functional-unit (FU) scans yield 979 genome-wide-significant leads, and diagnosis-weighted composites recapitulate genome-wide FU signal in FinnGen and MVP. **a**, Lead variants from all FU scans in 423,955 UK Biobank participants, ordered by SNP heritability. Dot area scales with *−* log_10_ *P* . Open rings mark leads in the major histocompatibility complex, and the BMI FU is shown translucently because of its high lead density. Row annnotations give lead counts and LDSC summary statistics. **b**, Genome-wide reconstructed associations for the cardiometabolic and atopic–immune airway FUs in FinnGen R12 and MVP R4 Europeans. Each track is a diagnosis-GWAS composite weighted by chromosome-held-out UK Biobank FU weights and calibrated on control variants. It is not an FU scan performed in the external cohort. Coloured points mark UK Biobank FU leads carried by that cohort. The dashed 5 *×* 10*^−^*^8^ line is a reference on The calibrated composite *P* value. Fig. 3f,g and Extended Data Fig. 6 show lead-level concordance.

**Extended Data Fig. 5.**
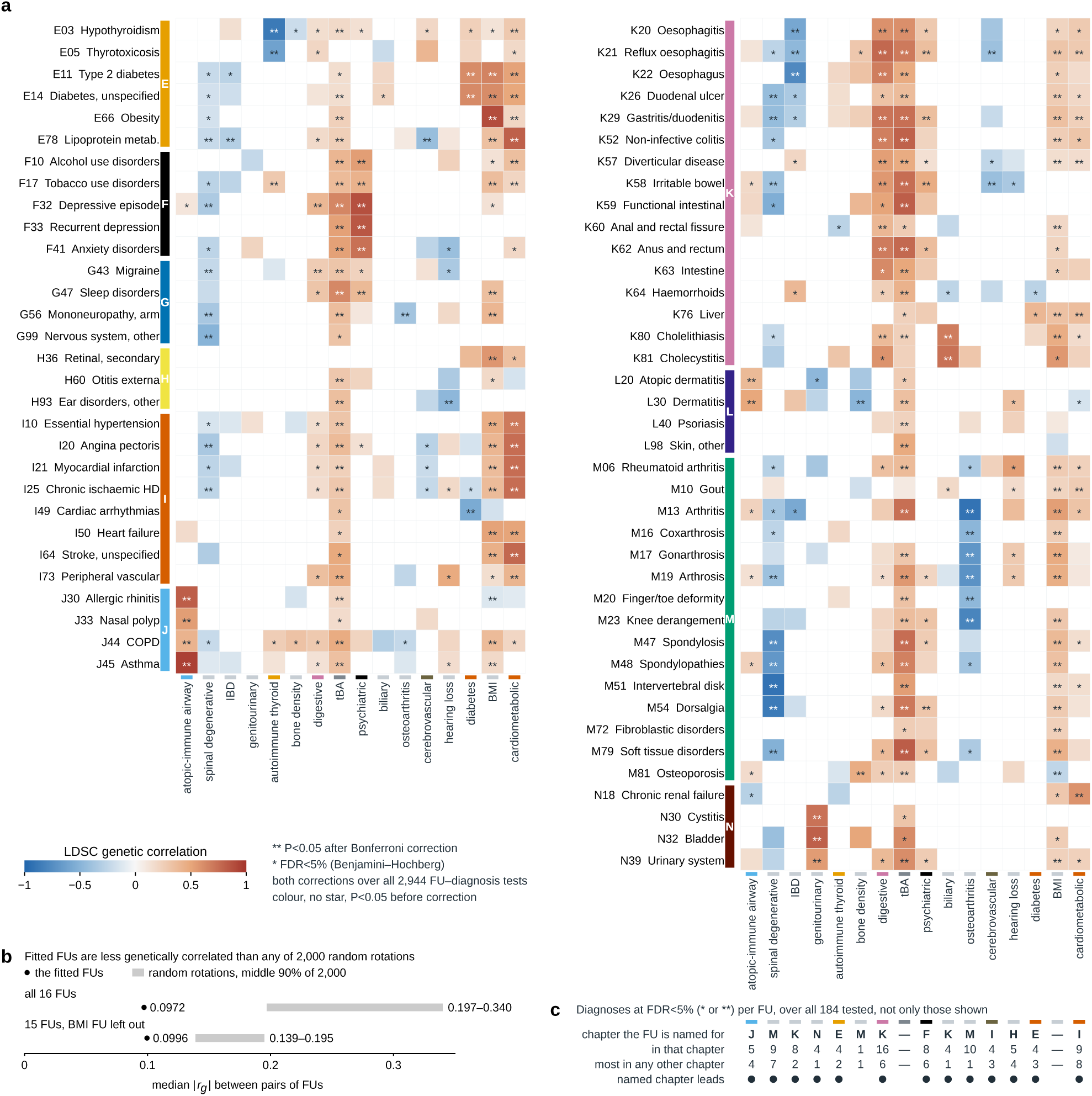
Functional-unit (FU) genetic correlations align with their clinical domains and are lower than expected after random rotation. **a**, LDSC genetic correlations between the 16 FU scans and 232 diagnosis scans, 184 of which gave estimable correlations. Multiple-testing correction and the counts in c use all 2,944 estimable cells, and the heat map shows the 69 diagnoses with at least one Bonferroni-significant correlation. Colour encodes *r_g_* for nominally significant cells, with asterisks marking 5% FDR and Bonferroni significance. **b**, Rotation control for pairwise FU genetic correlation. Dots show the median absolute *r_g_* among the fitted FUs, with and without the BMI FU. Bars show the middle 90% across 2,000 random orthonormal rotations. Both observed medians lie below all rotated values. **c**, For each FU, counts of FDR-significant FU–diagnosis correlations in the ICD-10 chapter named by that FU and in the strongest alternative chapter. Thirteen of the 14 chapter-named FUs have more significant correlations in their named chapter. BMI and tBA have no named chapter.

**Extended Data Fig. 6.**
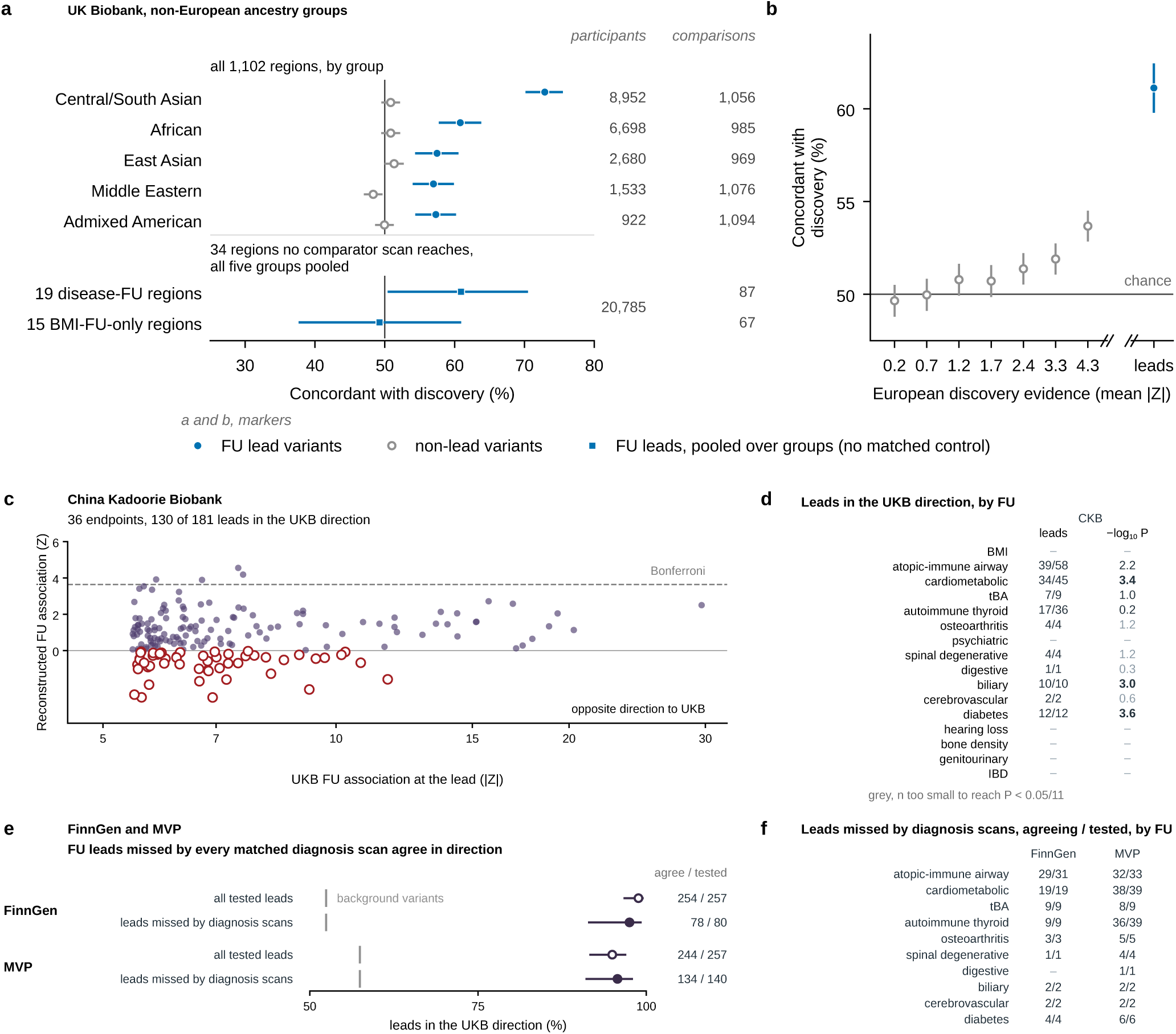
Functional-unit (FU) genetic direction is preserved across UK Biobank ancestry groups annod in diagnosis-weighted composites from external cohorts. **a**, Directional concordance of FU region leads in five non-European UK Biobank genetic-ancestry groups, compared with frequency-matched control variants. Squares mark the 34 regions missed by the specified comparators. **b**, Concordance of non-lead variants binned by European *|Z|*, with FU leads shown beyond the axis break. **c**, Reconstructed FU association at 181 tested leads in China Kadoorie Biobank (CKB). Positive *Z ind*icates agreement with UK Biobank. **d**, CKB agreement by FU with exact binomial tests against 0.5. CKB is exploratory and uses 36 matched diagnoses. Its autoimmune-thyroid composite does not represent that FU because no CKB thyroid endpoint reaches 500 cases. **e**, Directional agreement in FinnGen and MVP for all tested FU leads and for the subset at which no matched UK Biobank diagnosis reaches *P <* 5 *×* 10*^−^*^8^. Grey ticks show matched-background rates. **f**, Agreeing over tested leads in that missed-diagnosis subset, by FU. External reconstructed associations are weighted diagnosis composites, not second measurements of the FU phenotype. Error bars in a, b and e are 95% Wilson intervals.

**Extended Data Fig. 7.**
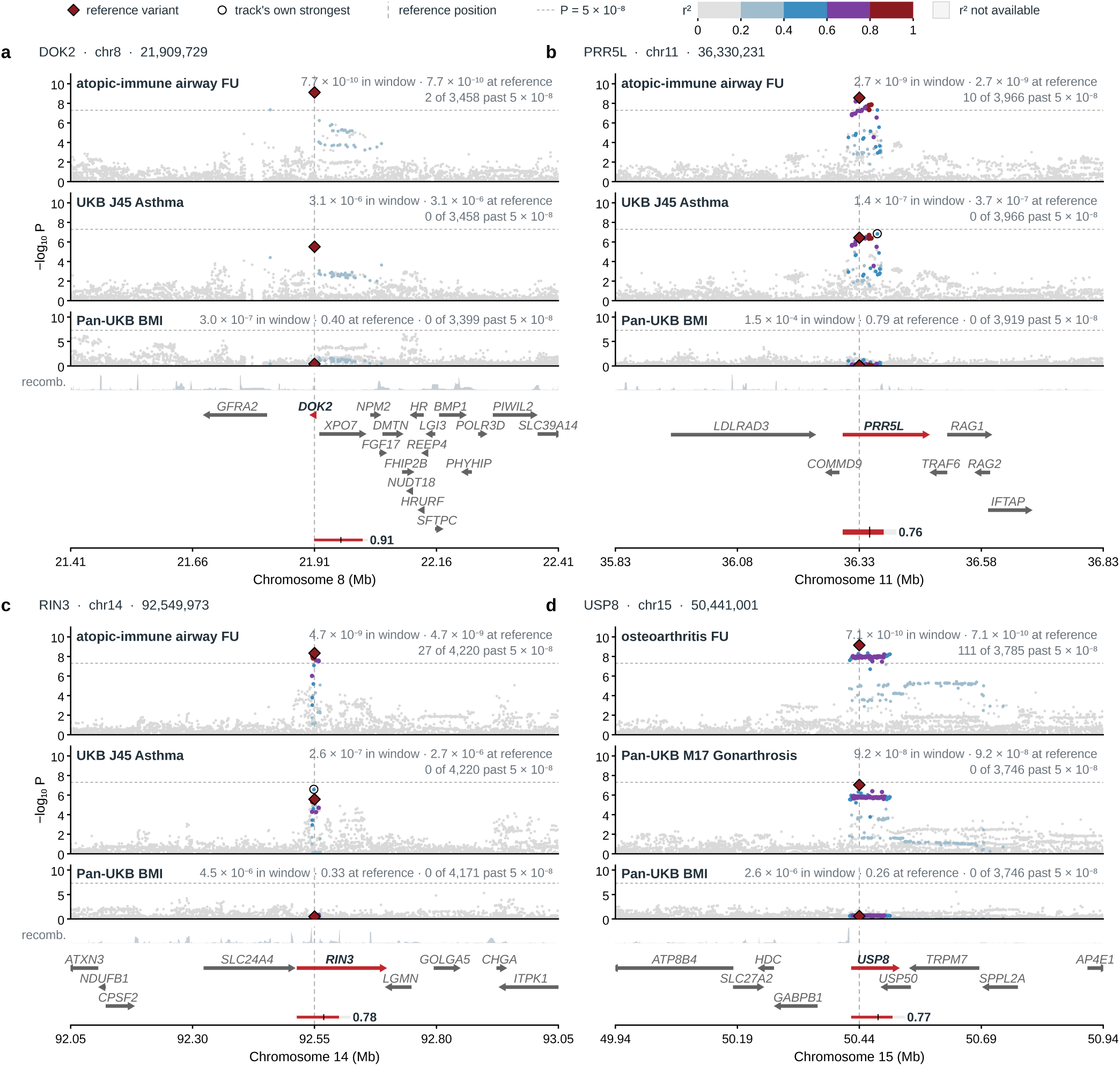
Four comparator-unreached regions with locus-to-gene scores of at least 0.5 show stronger FU than linked-diagnosis association. The four regions are selected by the stated L2G rule from the 34 regions of Fig. 4. each panel spans *±*500 kb and shows the FU scan, the linked diagnosis with the smallest regional *P* , Pan-UK Biobank BMI, recombination rate and protein-coding genes. **a**, Atopic–immune airway FU at the *DOK2* -labelled region. **b**, Atopic–immune airway FU at *PRR5L*. **c**, Atopic–immune airway FU at *RIN3*. **d**, Osteoarthritis FU at *USP8*. Colours show *r*^2^ to the regional reference variant and open symbols mark track-specific minima. FU and diagnosis scans are not independent. Gene names are positional labels rather than causal assignments.

**Extended Data Fig. 8.**
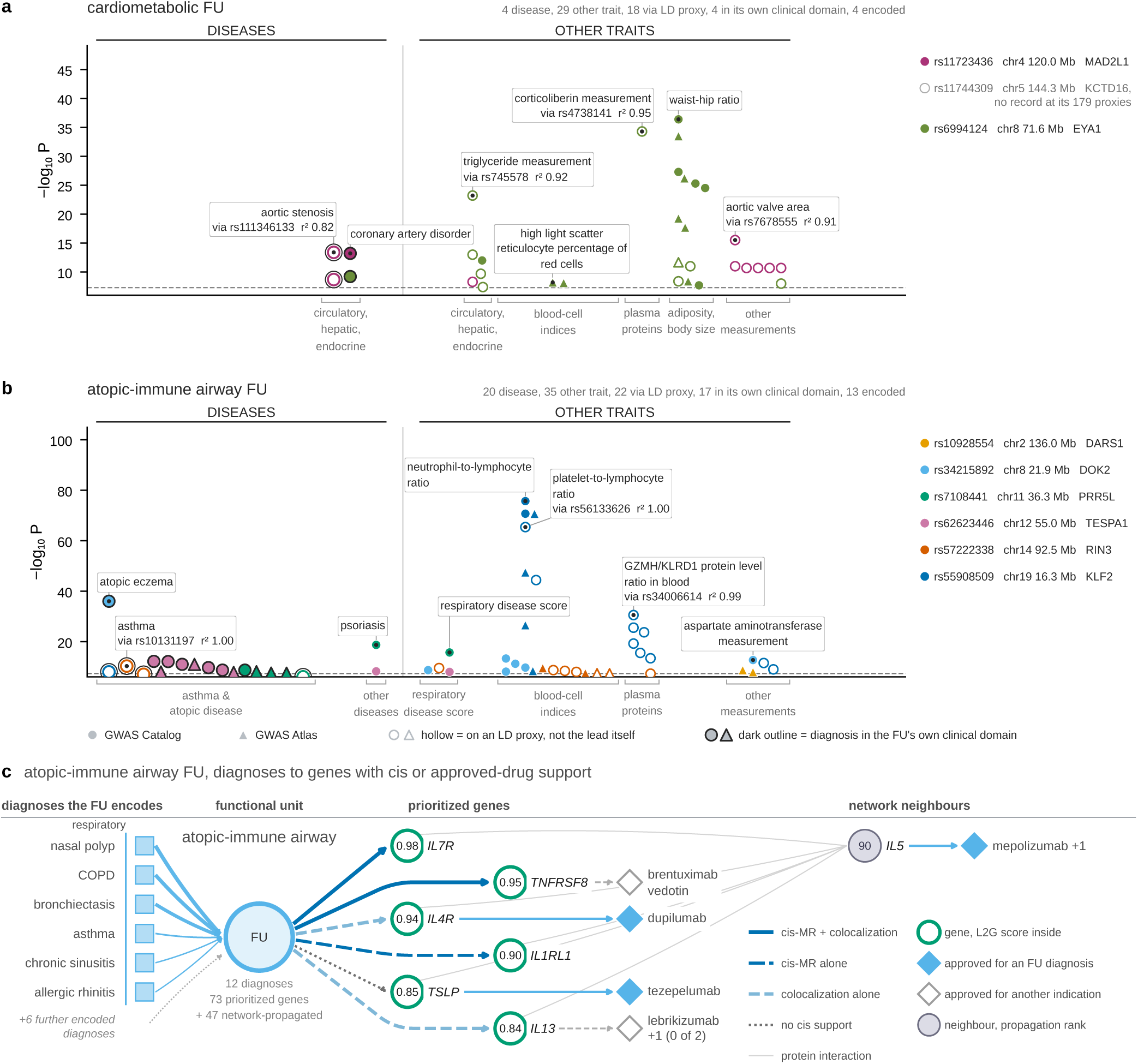
FU-specific leads carry clinically related catalogue associations and connect the atopic– immune airway FU to molecular and therapeutic evidence. **a**, **b**, GWAS Catalog and GWAS Atlas associations at the three cardiometabolic and six atopic–immune airway leads from Fig. 4b or at proxies with *r*^2^ *≥* 0.8. The display contains 88 associations, 85 at *P ≤* 5 *×* 10*^−^*^8^. Of 24 disease associations, 21 fall in the carrying FU’s clinical domain and 17 are diagnoses encoded by that FU. Catalogue retrieval is not exhaustive, so these records are a lower bound on external trait support. **c**, Atopic–immune airway evidence map drawn with the same logic as Fig. 5e. Six MR-APSS-supported respiratory diagnoses connect to genes selected by *cis* molecular evidence or by an approved drug for an encoded diagnosis. The display rule was applied to 25 of the FU’s 120 prioritized and propagated genes, and seven of the eight that meet it are shown. The panel is illustrative and is not an enrichment estimate.

**Extended Data Fig. 9.**
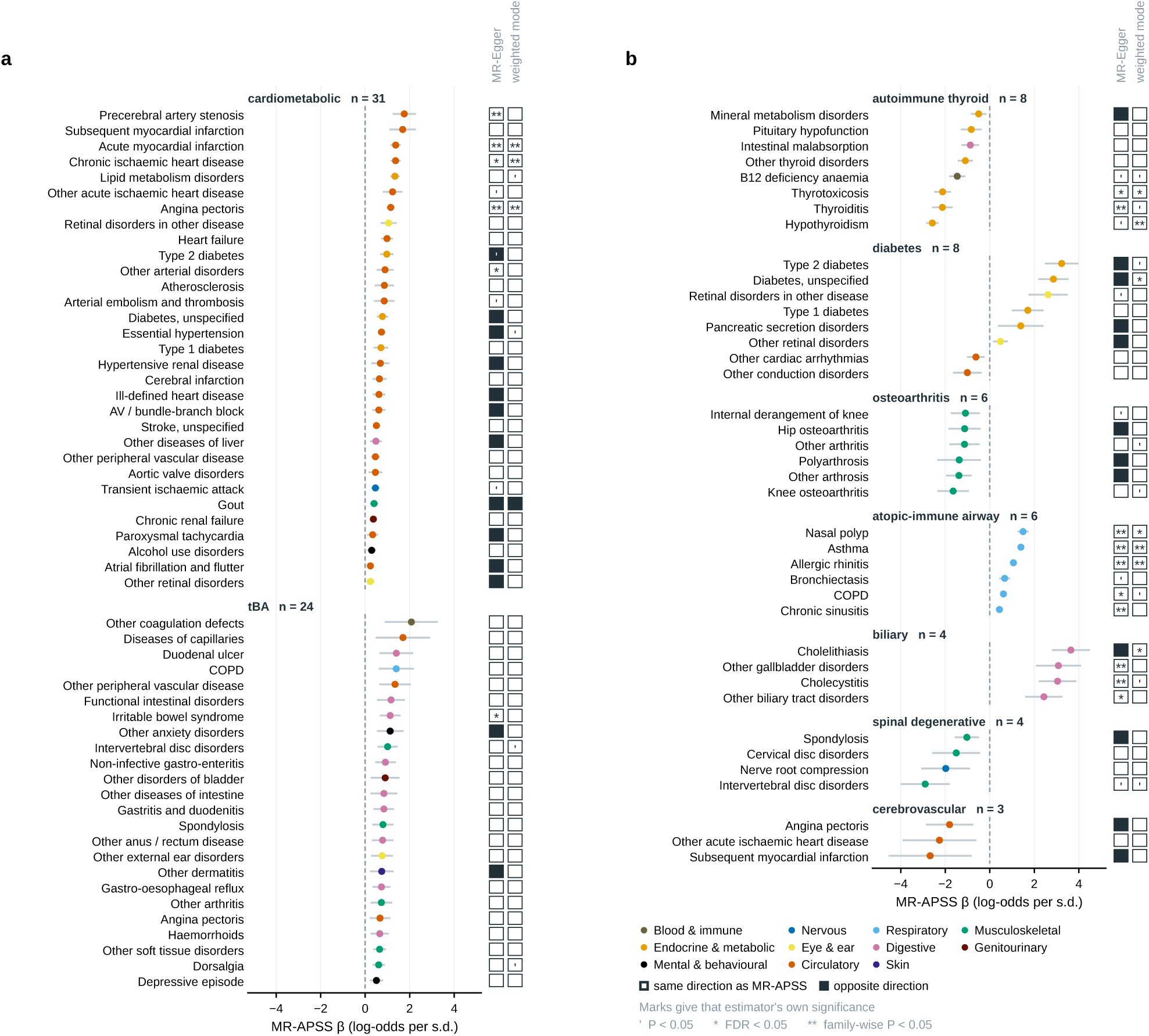
Genetic effects across linked diagnoses are directionally coherent within nine disease functional units (FUs). **a**, **b**, MR-APSS estimates for 94 FU–diagnosis associations passing 5% FDR within the single 804-pair family, shown as one continued list. Points and bars are MR-APSS estimates with 95% confidence intervals, in log-odds of the diagnosis per s.d. of the FU. Adjacent cells give MR-Egger and weighted-mode sensitivity estimates. White cells agree in direction with MR-APSS and black cells disagree. Primes and asterisks mark nominal, 5% FDR and Bonferroni significance for each estimator, with MR-Egger and weighted mode each corrected over its 587 estimable pairs. MR-Egger agrees in direction for 69 of 94 associations and weighted mode for 93 of 94. The linked diagnoses contribute to FU construction and the GWAS share participants, so these estimates describe genetic coherence within the defining clinical profiles rather than a causal effect of an FU on its constituent diseases. The 40 BMI-FU positive-control pairs are not drawn. With the Pan-UK Biobank BMI scan and the GIANT–UK Biobank BMI meta-analysis as exposures, MR-APSS estimates on their 40 diagnoses agree in sign with the BMI-FU estimates for all 40, with through-origin slopes against them of 0.976 and 1.016, a descriptive calibration and not an independent replication.

## Acknowledgements.

This research has been conducted using the UK Biobank Resource under Application Number 21988.

## Declarations

### Data availability

UK Biobank participant-level data are available to approved researchers through the UK Biobank access process. Data generated in this study and the analysis code will be made available upon publication. The external-cohort test used public summary statistics only. FinnGen release R12 summary statistics are distributed by the FinnGen consortium [29] at https://www.finngen.fi/en/access_results, Million Veteran Program release 4 European summary statistics are available through dbGaP under accession phs002453.v1.p1 [30], and China Kadoorie Biobank summary statistics come from the CKB PheWeb release at https://pheweb.ckbiobank.org described by Walters et al. [31]. No participant-level data were obtained from any of these three cohorts.

### Code availability

Analysis code and figure-generation scripts will be released upon publication.

## Supplementary Information

**Supplementary Fig. 1.**
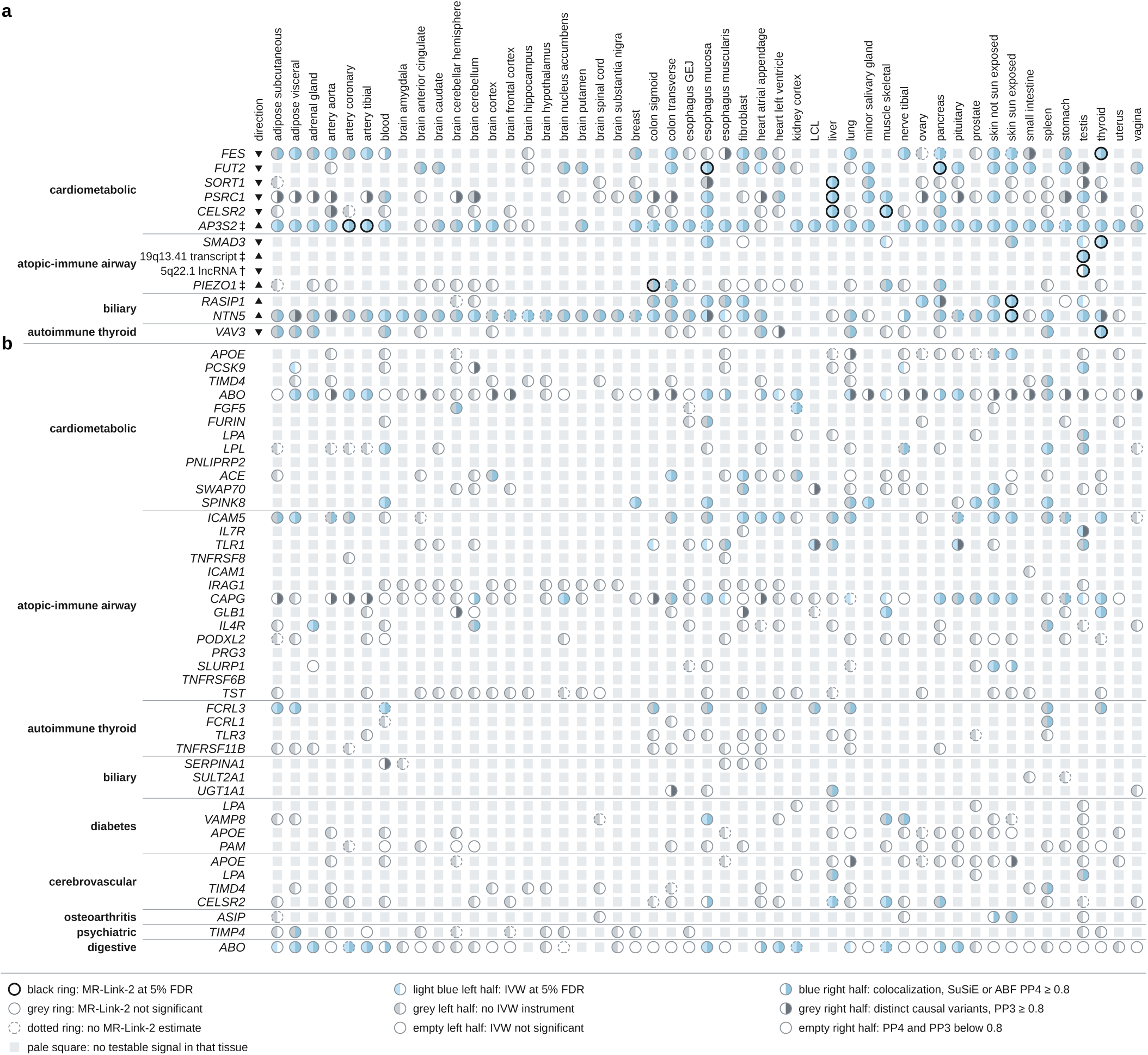
In 49 GTEx v8 tissues, MR-Link-2 and colocalization agree for 13 functional unit (FU)–gene pairs over four FUs, and eleven of them colocalize in more tissues than the one or two where MR-Link-2 passes, so the passing tissue need not be where the gene acts. Each circle is one gene in one tissue with a testable *cis* signal (smallest *P ≤* 5 *×* 10*^−^*^5^), read with the key below. Pale squares and dotted rings are absences, not negatives. MR-Link-2 and the inverse-variance-weighted (IVW) comparator are corrected at a 5% false-discovery rate (FDR) within each FU and tissue. Only one IVW pass here rests on two instruments, and it does not colocalize, so IVW selects no row. Colocalization uses the SuSiE result where SuSiE returned a credible-set pair and the approximate-Bayes-factor (ABF) result otherwise, never the larger of the two. Unlike a plasma protein assay, a transcript readout does not depend on reagent binding. Tissues are alphabetical, not ranked. Blood is whole blood, LCL a lymphoblastoid cell line and GEJ the gastroesophageal junction. **a**, The 13 pairs with a black ring and a blue right half in the same tissue, 16 FU–gene–tissue combinations, two of them through ABF. Triangles give the direction of the MR-Link-2 estimate, upward for more expression and a higher mortality-aligned FU value. *†*, an unnamed transcript (ENSG00000248428) testable in testis alone, where the MR-Link-2 pleiotropy term is significant, so it is not read as evidence for a gene. *‡*, the FU scan does not reach 5 *×* 10*^−^*^8^ in this window. Four of the 48 plasma-board pairs (*CELSR2*, *FES*, *SMAD3* and *SORT1*) are among the 13. For *SMAD3*, of the atopic–immune airway FU, the thyroid MR-Link-2 estimate points opposite to its UK Biobank inverse-variance-weighted plasma estimate, its only passing plasma estimate. **b**, The other 44 FU–gene pairs of the plasma board, rows the plasma analysis selected, drawn for comparison. *IRAG1* was formerly *MRVI1*.

**Supplementary Fig. 2.**
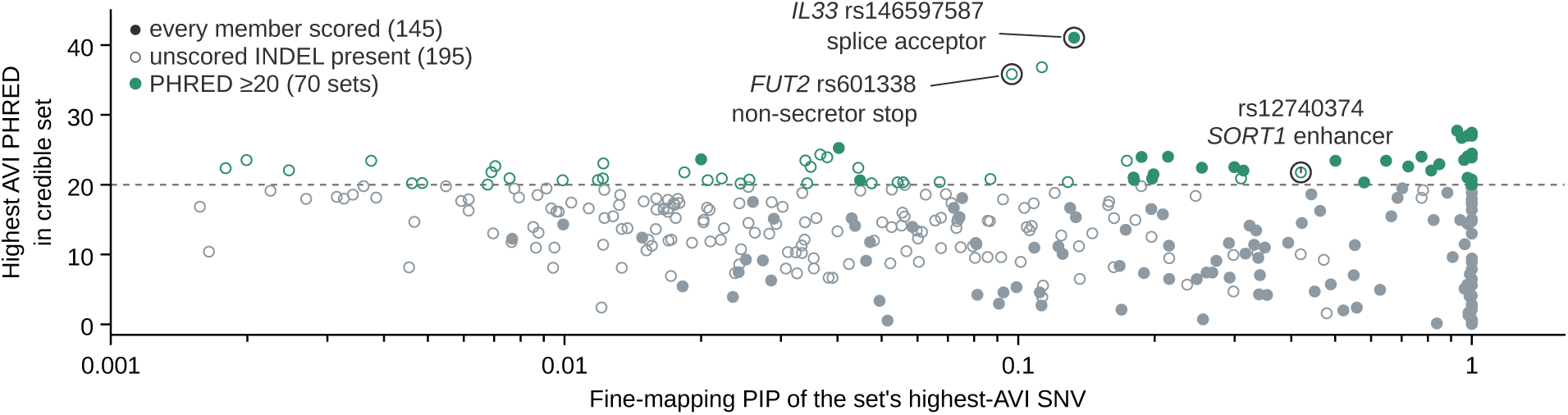
Sixty-nine of the 7,157 single-nucleotide variants in the credible sets of the 11 disease functional units (FUs) rank in the genome-wide top 1% of predicted variant impact, the rate the scale itself implies, and they place 70 of the 340 sets at that level. Highest AlphaGenome Atlas variant-impact (AVI) PHRED score among the scored SNVs of each of the 340 credible sets of the 11 disease FUs, against that SNV’s fine-mapping posterior inclusion probability (PIP, logarithmic axis). Green, 20 or more, the genome-wide top 1% of SNVs (70 sets). Filled, every member scored (145). Hollow, an unscored insertion or deletion is present (195) and a higher-scoring member could be missing. Points sit at their exact coordinates, so most touch a neighbour (235 of 340), and 29 sets stack at a PIP of 0.999 or more. Ringed, three alleles with published functional evidence [18–20], the *IL33* splice acceptor rs146597587 (PHRED 41.1, PIP 0.13, fourth of nine members), the *FUT2* non-secretor stop rs601338 (35.8, 0.10, first of 35) and the *SORT1* enhancer rs12740374 in the *CELSR2* 3*^′^* untranslated region (21.8, 0.42, first of six). AVI scores alleles and assigns no gene, tissue, direction or causal probability. Sixty-nine of 7,157 is 0.96%, so the 70 sets are not an enrichment, and the sets can outnumber the variants because five alleles lead a set in each of two FUs. The tissue expression readout (Supplementary Fig. 1) supports no *IL33* pair, and its *FUT2* pair lies on the cardiometabolic FU, not the digestive FU where rs601338 was fine-mapped, so neither allele gains a direction on the FU where it was fine-mapped.

